# Goal Attainment Scale light captures clinically meaningful changes in Adolescents and Adults with Spinal Muscular Atrophy Treated with Risdiplam

**DOI:** 10.64898/2026.08.02.26359391

**Authors:** Nancy Carolina Ñungo Garzón, Karolina Aragon-Gawinska, Inmaculada Pitarch Castellano, Teresa Sevilla, David Hervás, Juan F Vázquez-Costa

## Abstract

**Introduction/Aims:** To evaluate the usefulness of the Goal Attainment Scale (GAS) light for assessing response to risdiplam in patients with SMA aged ≥15 years.

**Methods:** In this population-based, longitudinal, ambispective study, patients were evaluated before and at 12 and 24 months after risdiplam initiation using motor scales (SMA Functional Composite Score Revised [SMA-FCR]), pinch strength (MyoPinch), functional scales (EK2, ALSFRS-R), patient and clinician global impression of change (PGIC and CGIC), and GAS light. Longitudinal changes were assessed using linear mixed-effects models. The minimal detectable change (MDC) and minimal clinically important change (MCIC) of GAS light were calculated.

**Results:** Forty-four patients (median age 32 years; 56.8% female) were included: 31.8% non-sitters, 56.8% sitters, and 11.4% walkers. GAS light priorities differed across functional subgroups, with patients prioritising moderately affected domains. After 24 months of risdiplam treatment, motor outcomes showed non-significant improvements in walkers, whereas functional scales improved significantly only in non-sitters. In contrast, GAS light detected significant, increasing improvements across all functional subgroups. The MCIC and MDC for GAS light were 6.5 and 10.65 points, respectively. According to the CGIC, 58% of patients improved slightly, 29% remained stable, and 13% worsened slightly at 24 months. Using the MCIC threshold, 64.5% achieved clinically meaningful goal improvement.

**Discussion:** GAS light is a feasible, sensitive, patient-centred tool that may complement standardised outcome measures when evaluating treatment response in adults with SMA. These findings further support risdiplam as a valuable therapeutic option in this population.

## INTRODUCTION

5q spinal muscular atrophy (SMA) is an autosomal recessive neuromuscular disorder caused by a deficiency of the survival motor neuron (SMN) protein due to biallelic variants in the *SMN1* gene. It is characterised by a progressive degeneration of anterior horn cells, leading to muscle weakness, atrophy, and functional impairment of variable severity. Historically considered a paediatric disease, advances in multidisciplinary care and the advent of disease-modifying therapies (DMTs) have significantly improved survival, resulting in a growing population of adults living with SMA[1].

DMTs dramatically change the natural history of SMA when patients are treated before symptom onset or early after. However, in adolescent and adult SMA patients, goals and long-term responses to DMTs differ considerably from those in paediatric populations[2–4]. Rather than large motor gains, treatment goals and achievements usually consist of preservation of independence, reduction of fatigue, or mild improvements in some specific daily activities[2–4]. These outcomes are not always adequately captured by traditional motor scales such as the Hammersmith Functional Motor Scale Expanded (HFMSE) and the Revised Upper Limb Module (RULM), which were developed for paediatric patients but show considerable limitations in adults, including floor and ceiling effects and limited responsiveness to change[3, 5–8]. Also, functional scales validated in adult SMA patients, such as the Amyotrophic Lateral Sclerosis Functional Rating Scale-Revised (ALSFRS-R) or Egen Klassifikation 2 (EK2), fail to fully reflect the clinical heterogeneity of SMA presentation in adult patients[5, 9–12].

These limitations have driven increasing interest in patient-reported outcome measures (PROMs), which capture domains that matter most to individuals living with SMA, including fatigue, participation, respiratory function, and overall well-being[13–15].

However, many available PROMs lack robust validation (especially on responsiveness) in adult SMA and remain constrained by fixed-item structures.

Goal Attainment Scale light (GAS light) represents a personalised and patient-centred alternative to traditional PROMs. Through a structured collaboration between clinician and patient, individualised and functionally meaningful goals are defined and rated on a standardised five-point scale, enabling both personalisation and quantification of response.[16, 17] By defining success according to patient-prioritised outcomes, GAS has the potential to detect meaningful changes that conventional scales may overlook[6, 14, 18–20]. Casiraghi et al. recently showed that, while most of the goals selected in GAS overlapped with items included in existing motor scales, other items reflected personalised domains (such as activities of daily living, endurance, or functional mobility) that may not be captured by conventional motor assessments[18]. Moreover, Cintas et al. recently found that these patient-centred tools were more sensitive in detecting improvement in treated adult SMA patients than the standard Motor Function Measure - 32 items scale (MFM32)[21].

Thus, the growing use of DMTs in adult SMA, alongside the limitations of existing scales, raises the need for outcome measures that capture effectiveness in real-world settings. This population-based study examines the clinical value of GAS light for the longitudinal assessment of response to risdiplam treatment in adolescents and adults with SMA.

## METHODS

### Study Design and Participants

This was a population-based, longitudinal, ambispective study including patients who initiated treatment with risdiplam at the Neurology outpatient clinic of Hospital La Fe (Valencia, Spain).

#### Patient population

Currently, all SMA patients living in the Valencian Community (ca. 5 million inhabitants) are being treated and followed up at Hospital La Fe. For this study, we retrospectively reviewed all genetically confirmed SMA patients aged ≥15 years visited at Hospital La Fe and included those who initiated treatment with risdiplam in our centre between November 2020 and November 2025.

#### Variables

Demographic, clinical and genetic data were collected. Patients were classified according to SMA type, based on age at symptom onset and the highest motor milestone achieved. Participants were also categorised according to functional status as non-sitters (unable to maintain a seated position upright with the head erect for at least 3 seconds without using the arms or hands for support); sitters (able to sit independently and maintain a stable upright position for at least 3 seconds without support, but unable to walk at least 10 meters without support); and walkers (able to walk at least 10 meters without support).

Outcome measures were prospectively collected at baseline (right before treatment initiation) and at 12 and 24 months of follow-up, including motor (HFMSE, RULM and 6-Minute Walk Test - 6MWT) and functional scales (EK2, ALSFRS-R), together with the maximal thumb pinch strength of the dominant hand measured with MyoPinch in kilograms (kg). Details can be found as supplementary material. Additionally, the Patient - Global Impression of Change scale (PGIC) and the Clinician - Global Impression of Change scale (CGIC) using a 1 to 7 Likert scale were collected in both follow-up visits.

GAS light was administered at baseline in all patients. At least three individualised, functionally meaningful goals were collaboratively defined for each patient after a discussion between the patient and an expert neurologist (JFVC) using the SMART framework (Specific, Measurable, Achievable, Relevant, Time-bound). Goals were categorised into six functional regions (axial, bulbar, respiratory, upper limb, lower limb, and general stamina), which were further subdivided into functional domains according to individual patient tasks (Supplementary table 1). For each goal, patients together with the expert clinician scored the degree of baseline performance (-2 for no function, -1 for partial function) and rated its importance and the perceived probability of achievement in a 1 to 3 scale, using the GAS light worksheet and instructions developed by King’s College London (https://www.kcl.ac.uk/cicelysaunders/resources/toolkits/gas-overview).

The achievement of each goal was reassessed at 12 and 24 months by consensus of the clinicians and the patients, using the following scores: -2 when the goal did not improve or improved much less than expected; -1 when the goal was only partially achieved; 0 when the goal was achieved as expected; and +1 when the goal was achieved somewhat more, or much more (+2) than expected. Finally, when the patient did not perceive any changes, the value recorded at baseline function was maintained.

### Statistical analysis

All statistical analyses were performed using R software (version 4.5.1). Baseline characteristics were summarised using descriptive statistics. Continuous variables are reported as mean and standard deviation (SD) or median and interquartile range (IQR), as appropriate, and categorical variables as frequencies and percentages.

The Spinal Muscular Atrophy Functional Composite Revised (SMA-FCR) was calculated, following the method described by Pasternak et al.[4], as the unweighted average of three percentage scores: HFMSE, RULM, and 6MWT. Non-ambulant participants were assigned a value of 0 on the 6MWT.

To evaluate longitudinal changes over time, linear mixed-effects models were used for SMA-FCR, MyoPinch strength, EK2, ALSFRS-R, and GAS light scores. Time was modelled as a fixed effect (visits: baseline, 12 months, and 24 months), functional status (non-sitter, sitter, walker) and its interaction with time were included as fixed effects to explore differential trajectories across subgroups; and age and sex were included as covariates. A random intercept for each patient was included to account for within-subject correlation across repeated measurements.

To analyse GAS light goal-setting patterns, relative frequencies of importance ratings were calculated and used to construct importance matrices stratified by functional status and functional region, as well as by functional status and functional domain. Mean probabilities of goal achievement and baseline performance were computed and summarised in probability matrices according to functional subgroup and region.

We also assessed the Minimal Detectable Change (MDC) and the Minimal Clinically Important Change (MCIC) of the GAS light. The MDC was estimated according to the method proposed by Schmitt and Di Fabio[22], using the formula MDC = 1.96 × √2 × SEM, where the Standard Error of Measurement (SEM) was calculated as SEM = SD × √(1 − r), with SD being the standard deviation of the baseline GAS light score in the cohort, and *r* the instrument’s reliability coefficient. A reliability value of r = 0.88 was used, corresponding to the absolute-agreement intraclass correlation coefficient (ICC[A,1] = 0.88; 95% CI: 0.80–0.93) previously reported for the GAS light[23].

The MCIC was determined using the CGIC as the anchor. Receiver Operating Characteristic (ROC) analysis was performed using minimally perceived improvement (CGIC: ‘minimal improve’ vs. ‘unchanged’) as the classification variable and the change in GAS light score between baseline and follow-up as the continuous variable. The optimal cut-off point was identified as the one maximising the Youden index (sensitivity + specificity − 1) [24].

#### Ethics

All patients signed informed consent for participation in CUIDAME registry (NCT07231549). The study was approved by the Ethics Committee for Biomedical Research of Hospital La Fe (2019-027-1) in accordance with the Declaration of Helsinki.

## RESULTS

### Study population

During the study period, 47 patients started risdiplam treatment, but 3 patients were excluded from the study because they lacked the baseline GAS light. The study population comprised 44 patients (56.8% female), with a median age of 32 years (IQR: 24-46.75; range 15–73). Most patients were non-ambulant (31.8% non-sitters, 56.8% sitters vs 11.4% walkers) and 59.1% were treatment naïve while 40.9% had received prior nusinersen treatment. Other clinical characteristics can be found in Table 1.

**Table 1.** Demographic and clinical characteristics at baseline, stratified by functional status. Values are expressed as median [interquartile range (IQR)] for continuous variables and as number (percentage) for categorical variables, unless otherwise specified. Functional status was defined as non-sitter (unable to maintain a seated position upright with the head erect for at least 3 seconds without using the arms or hands for support), sitter (able to sit independently but unable to walk for ≥10 m unassisted), and walker (able to walk for ≥10 m without assistive devices). The 6-Minute Walk Test (6MWT) is applicable only to ambulant patients and was therefore not assessed in non-sitters or sitters (NA, not applicable). **Abbreviations:** 6MWT, Six-Minute Walk Test; ALSFRS-R, Amyotrophic Lateral Sclerosis Functional Rating Scale-Revised; BMI, body mass index; EK2, Egen Klassifikation 2; HFMSE, Hammersmith Functional Motor Scale Expanded; IQR, interquartile range; RULM, Revised Upper Limb Module; SMA, spinal muscular atrophy.

| Table 1. Demographic and clinical characteristics |  |  |  |  |
| --- | --- | --- | --- | --- |
| Functional State |  | Non sitters | Sitters | Walkers |
| Patients, n (%) |  | 14 (31.8%) | 25 (56.8%) | 5 (11.4%) |
| Median Age, years (range) |  | 31 (range 15-67) | 32 (range 16-73) | 26 (range 17–63) |
| Sex | Female | 8 (57.1%) | 14 (56%) | 3 (60%) |
|  | Male | 6 (42.9%) | 11 (44%) | 2 (40%) |
| SMA type | 2 | 11 (78.6%) | 9 (36%) | 0 (0%) |
|  | 3 | 3 (21.4%) | 16 (64%) | 5 (100%) |
| Scoliosis |  | 14 (100%) | 18 (72%) | 0 (0%) |
| Non-invasive ventilation (NIV) |  | 8 (57.1%) | 4 (16%) | 0 (0%) |
| Previous nusinersen treatment |  | 5 (35.7%) | 12 (48%) | 1 (20%) |
| HFMSE Baseline [IQR] |  | 0 | 5 (2-10) | 60 (45 – 61) |
| RULM dominant hand Baseline [IQR] |  | 1 (0–6.25) | 17 (16–24) | 37 (36–37) |
| 6MWT (m) [IQR] |  | NA | NA | 438 (296-512) |
| EK2 baseline [IQR] |  | 28 (23-30.8) | 14 (8-16.2) | 2 (1-4) |
| ALSFRS-R baseline median [IQR] |  | 22.5 (18.5-24) | 29 (28-33) | 43 (40-44) |

### GAS light Goals

Patients established 157 goals. Overall, upper limbs were the most frequently selected region (42%), but therapeutic priorities differed considerably according to functional status (Figure 1A and B, Supplementary Table 2). Most goals (81%) were related to functional abilities in which patients retained at least partial capacity, whereas only 19% addressed domains characterised by complete functional loss, mostly in non-sitter patients (Supplementary Figure 1). In addition, more goals, 138 (87.9%) targeted improvement and only 19 (12.1%) targeted stabilisation. Stabilisation goals focused on upper limb function (7, 36.8%), respiratory function (7, 36.8%), bulbar function (2, 10.5%), and axial, lower limb, and general stamina (1, 5.2%, each). Improvement goals were distributed as follows: upper limbs 42.7% (59), lower limbs 15.2% (21), respiratory 13% (18), axial 11.6% (16), generalised stamina 10% (14), and bulbar 7.2% (10). Finally, 78% of the goals were considered as either possible or probable to achieve with risdiplam (Supplementary Figure 2), indicating a pragmatic alignment between residual functional reserve and expectations of therapeutic benefit.

**Fig. 1.**
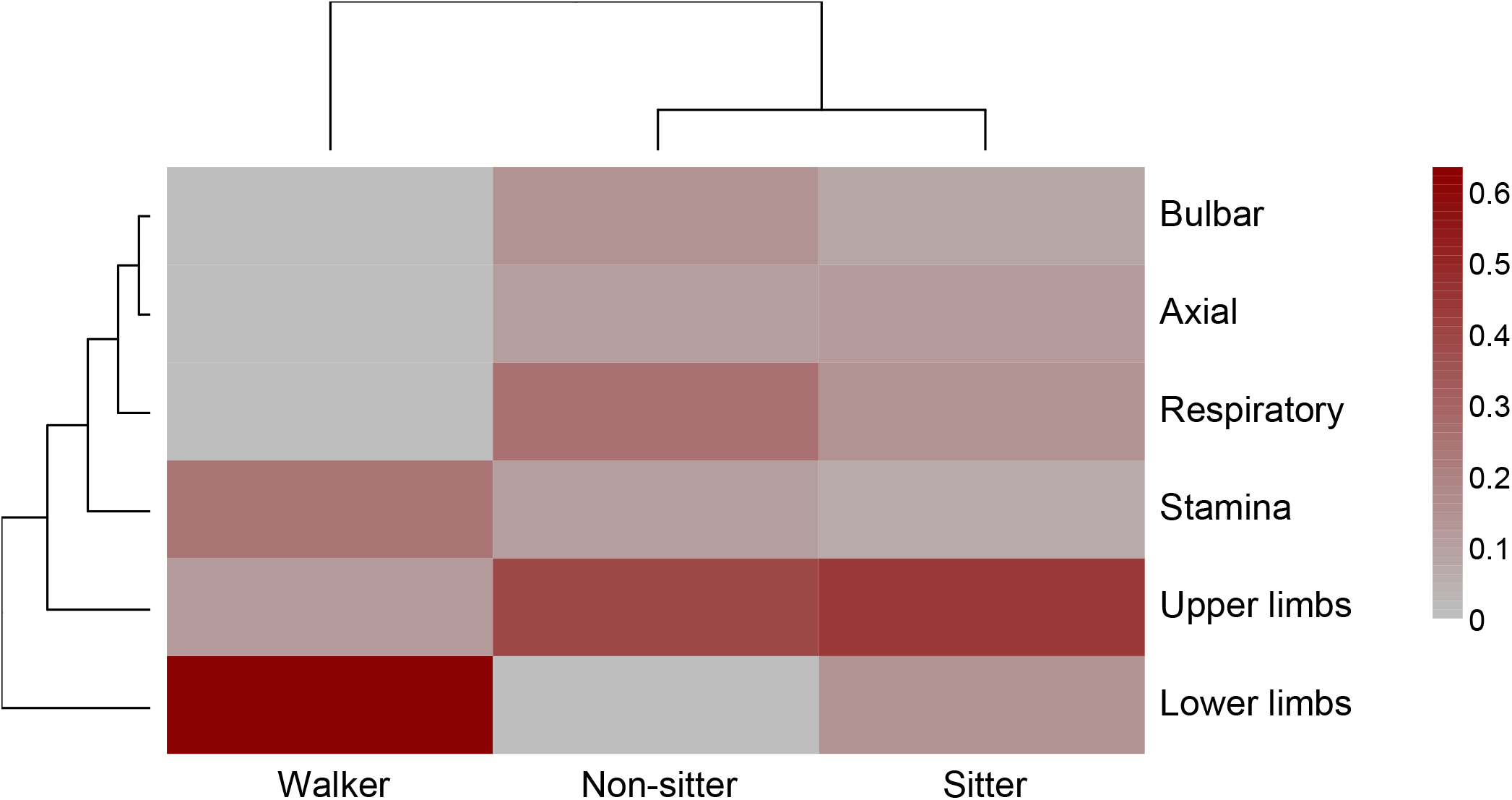
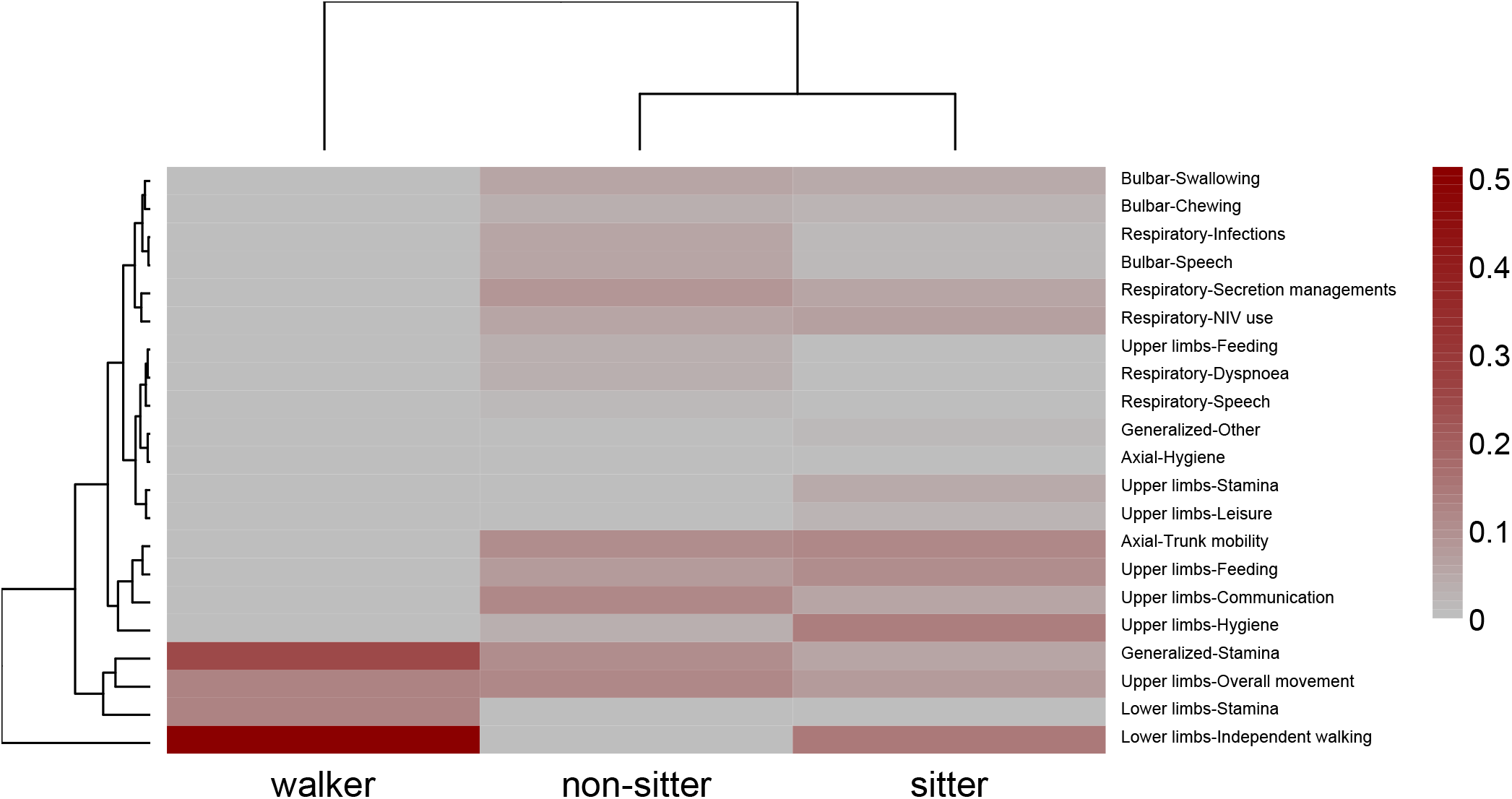
Weighted frequency of GAS light goals across functional regions (A) and region–domain combinations (B), according to baseline functional status. (A) Heatmap displaying the weighted frequency of goals set by patients in each functional region during the baseline GAS light interview, stratified by functional group (non-sitters, sitters, and walkers). (B) Heatmap displaying the weighted frequency of patient-set GAS light goals across the cross-classification of functional region and functional domain, stratified by the same functional groups. Colour intensity ranges from 0 (region/domain not prioritised) to the maximum relative importance within each panel. The pattern reveals a clinically coherent gradient aligned with residual functional capacity: upper limbs dominate in non-sitters and sitters, with dominant domains centred on independent mobility, hygiene, communication, and feeding; the respiratory region—particularly secretion management, non-invasive ventilation, and infection prevention—emerges as a differential priority among non-sitters; and lower-limb domains (independent mobility and stamina) are exclusively prioritised by walkers and a subset of sitters. **Abbreviations:** GAS, Goal Attainment Scale

Among non-sitters, upper limb function was the most frequently selected (93%) and important region for patients, followed by respiratory function. Upper limb–related domains primarily focused on preserving communication (sending messages), improving upper limb mobility (wheelchair control, transference, grasping objects), and feeding independence. Respiratory goals reflected the aim to improve secretion management and reduce respiratory infections and related hospitalisations. Avoiding the initiation of non-invasive ventilation (NIV) or reducing NIV dependence was also identified as a relevant objective in this subgroup. Overall, 96% of the goals targeted improvement and only 4% stabilisation (avoiding NIV initiation).

In sitters, upper limb function was also the most frequently selected region (92%) and the most important one, followed by respiratory. The most important domains for sitters were mainly related to regaining or maintaining independence in feeding, hygiene, stamina and mobility. Overall, 17.8% of goals targeted stabilisation, mainly in upper limbs and respiratory domains.

Among ambulatory participants, goals were predominantly related to lower-limb function (100%), followed by general stamina (60%). All goals were improvement-oriented, with independent walking representing the most frequently selected and important domain.

### Follow-up

Thirty-nine of the initial 44 patients completed at least 12 months of risdiplam treatment (1 patient died, 1 discontinued treatment early, 1 was lost to follow-up, and 2 had not yet reached the 12 months visit) and 31 reached the 24 months visit (1 additional discontinuation after 12 months; 7 not yet reached the 2 years follow up window).

### Motor Outcome Measures response

The SMA-FCR score remained stable in non-sitters and sitters and showed a minimal improvement in 3 out of 4 walkers (Figure 2A). The multivariable model, adjusting for age and sex, confirmed these results showing a small improvement in walkers (β = 0.868; 95% CI: −0.285 to 2.02; p=0.148) that did not reach statistical significance, possibly due to the small sample size (Supplementary Table 3).

**Fig. 2.**
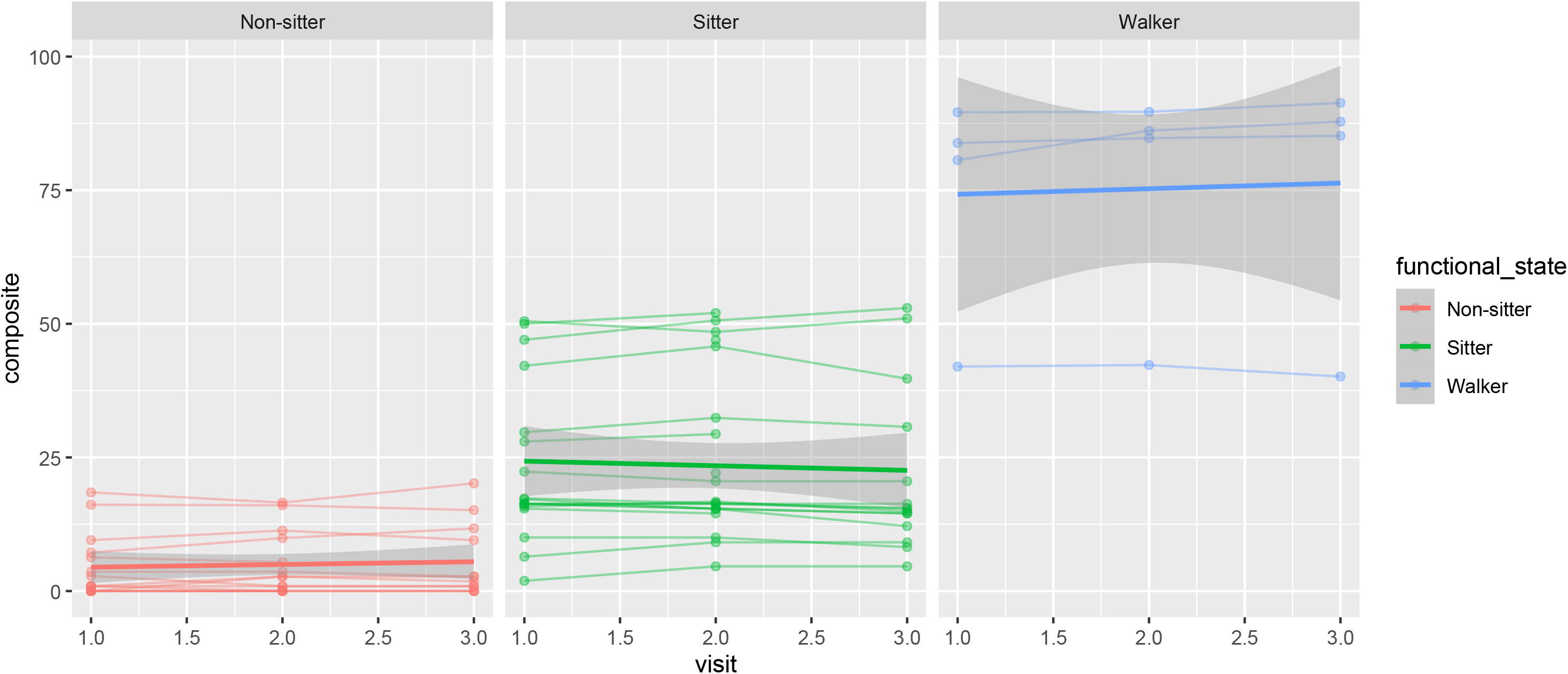
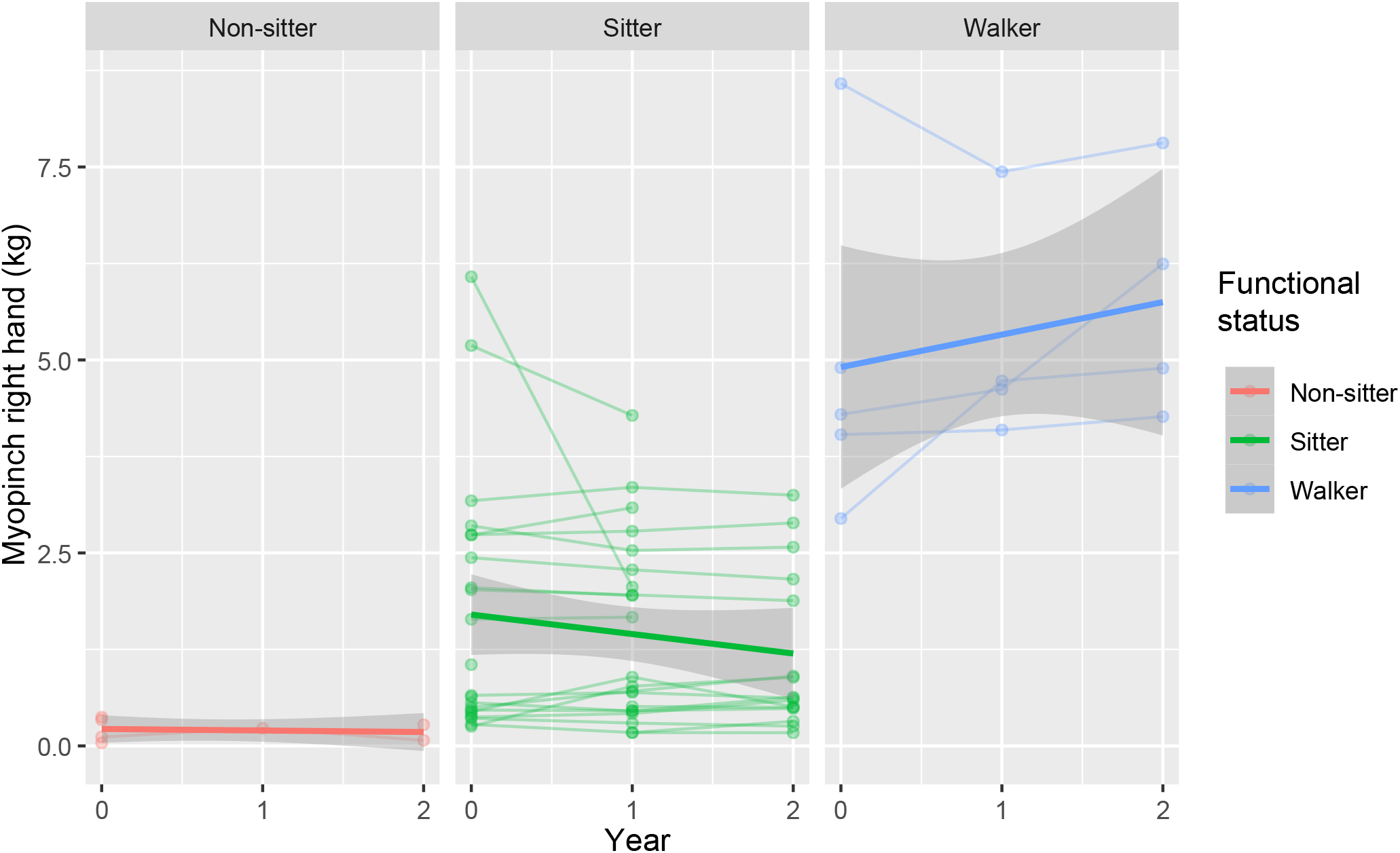
Longitudinal evolution of motor outcome measures across the two-year follow-up, stratified by baseline functional status. Boxplots displaying the distribution of (A) the Spinal Muscular Atrophy Functional Composite Revised (SMA-FCR) score and (B) dominant-hand pinch strength (MyoPinch, in kilograms-force, kg), at baseline (Year 0), at the 12-month follow-up (Year 1), and at the 24-month follow-up (Year 2), separately for non-sitters, sitters, and walkers. Pinch strength was measured using a calibrated MyoPinch dynamometer following a standardised protocol; the dominant hand was assessed in all patients. **Abbreviations:** kg, kilograms-force; SMA-FCR, Spinal Muscular Atrophy Functional Composite Revised.

Similarly, pinch strength of the dominant hand improved in 3 out of 4 walkers, but no consistent change was found in most sitters and non-sitters (Figure 2B). These results were confirmed in the multivariable model after adjusting for age and sex, where only a non-significant pinch increase in walkers (β = 0.471; 95% CI: −0.163 to 1.113; p=0.161) was appreciable (Supplementary Table 4).

### Functional Outcome Measures response

EK2 scores decreased in non-sitters and showed no appreciable changes in sitters (Figure 3A). The multivariable model confirmed this decrease over time in non-sitters (β = -1.365; 95% CI: -1.99 to -0.745; p<0.001). The sitter-visit interaction (β = 1.038; 95% CI: 0.256 to 1.826; p=0.012) indicated a different trajectory vs non-sitter, consistent with the stability observed in Figure 3A (Supplementary Table 5).

**Fig. 3.**
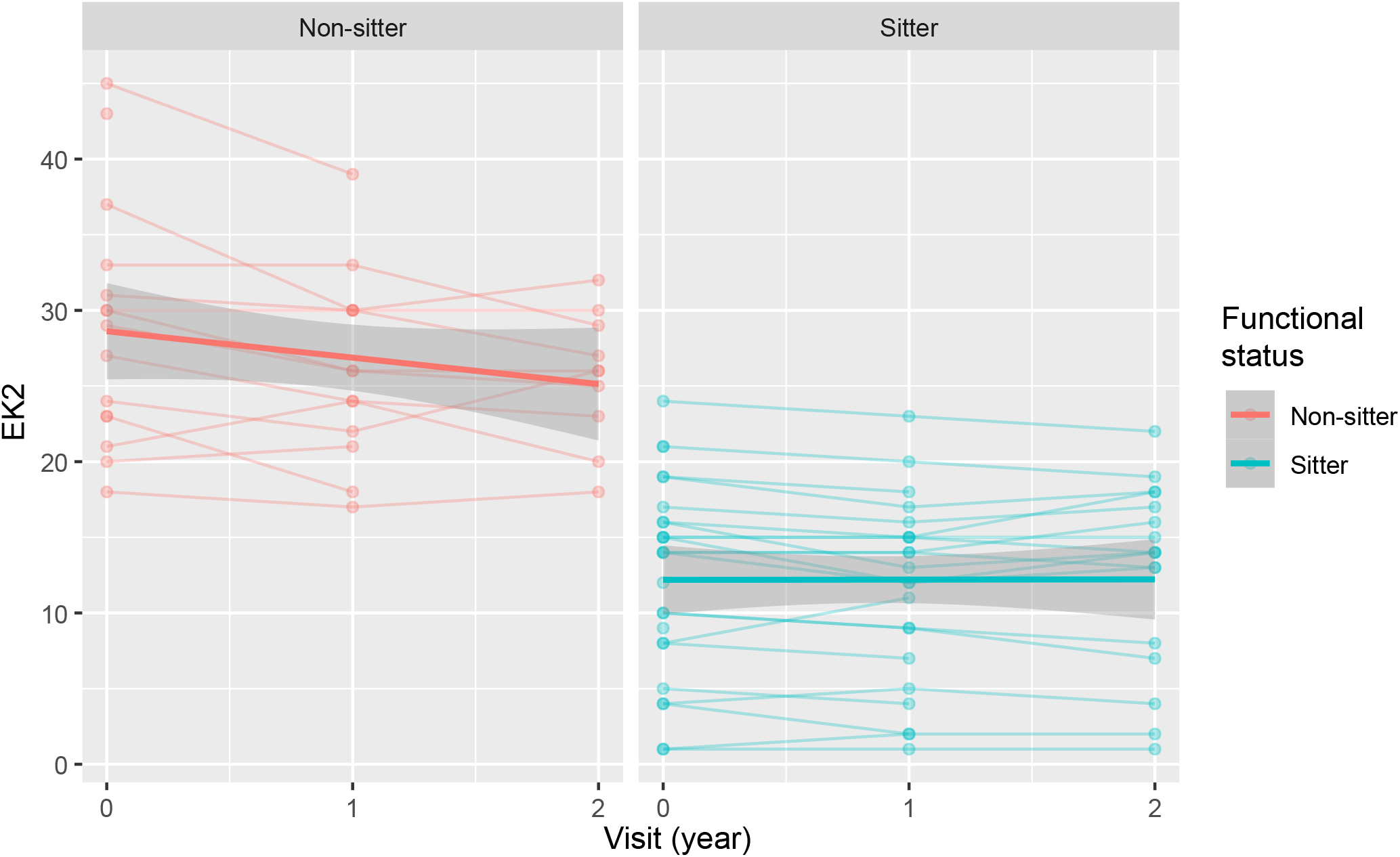
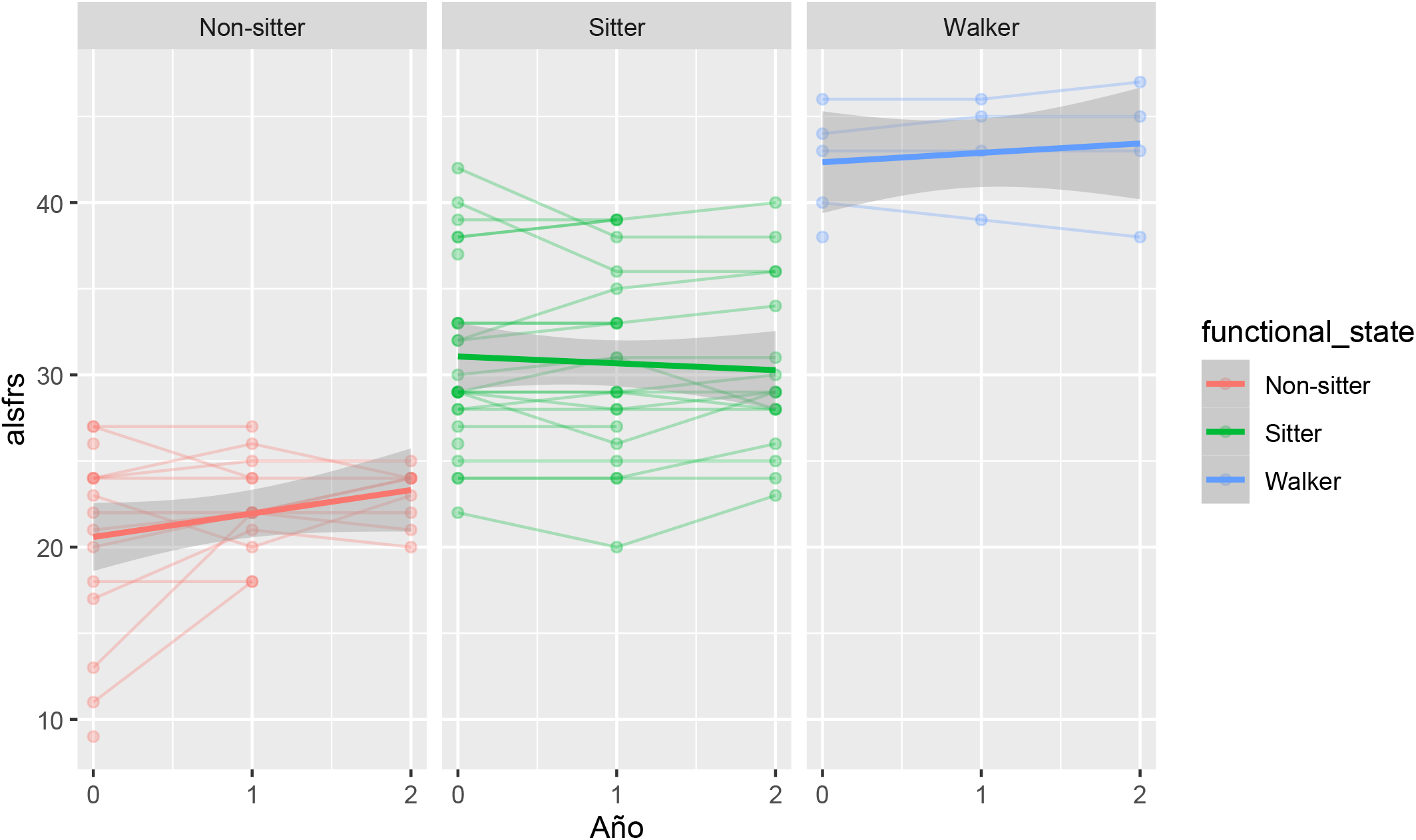
Longitudinal evolution of functional outcome measures across the two-year follow-up, stratified by baseline functional status. Boxplots displaying the distribution of (A) the Egen Klassifikation 2 (EK2) score and (B) the Amyotrophic Lateral Sclerosis Functional Rating Scale-Revised (ALSFRS-R) score, at baseline (Year 0), at the 12-month follow-up (Year 1), and at the 24-month follow-up (Year 2). EK2 is applicable only to non-ambulant patients and is therefore shown separately for non-sitters and sitters (Panel A), whereas ALSFRS-R is shown for non-sitters, sitters, and walkers (Panel B). Higher EK2 scores indicate greater disability, whereas higher ALSFRS-R scores indicate better functional status; therefore, a downward trajectory in EK2 and an upward trajectory in ALSFRS-R both reflect functional improvement. **Abbreviations:** ALSFRS-R, Amyotrophic Lateral Sclerosis Functional Rating Scale – Revised; EK2, Egen Klassifikation 2.

Similarly, the ALSFRS-R scores improved in non-sitters, but not in walkers or sitters (Figure 3B). The model confirmed a statistically significant improvement in non-sitters (β = 1.079; 95% CI: 0.431 to 1.734; p=0.002), but not in sitters or walkers (Supplementary Table 6).

### Global Impression of Change: Clinician and Patient Perspectives

According to the CGIC, 67.6% of patients were rated as slightly improved, 27% as stable and 5.4% as slightly worse at 12 months. At 24 months, 58% were rated as slightly improved, 29% as stable and 13% as slightly worse (Supplementary Figure 3A).

Similarly, the PGIC indicated that most patients perceived either improvement or stabilisation over the 2-year follow-up period (51.6% and 38.7%, respectively), whereas only 9.7% reported slight worsening. However, during the first 12 months, non-sitters and sitters tended to perceive greater improvements than those recognised by clinicians. This discrepancy was no longer evident at the 24-month follow-up (Supplementary Figure 3B).

### GAS light Changes with time

Of the 39 patients completing at least 12 months of risdiplam treatment, GAS light reassessment was available in 37 at 12 months and 31 at 24 months.

Median GAS light score change was 8.5 (3.2 in walkers, 8.5 in sitters, 11.1 in non-sitters) at 12 months, and 12.8 (8.75 walkers, 13.6 in sitters, 10.1 in non-sitters) at 24 months. The magnitude of improvement was similar across all phenotypes (Figure 4). The multivariable model confirmed that GAS light scores improved significantly over time after risdiplam treatment in all functional subgroups, independent of age and sex (Supplementary Table 7).

**Fig. 4.**
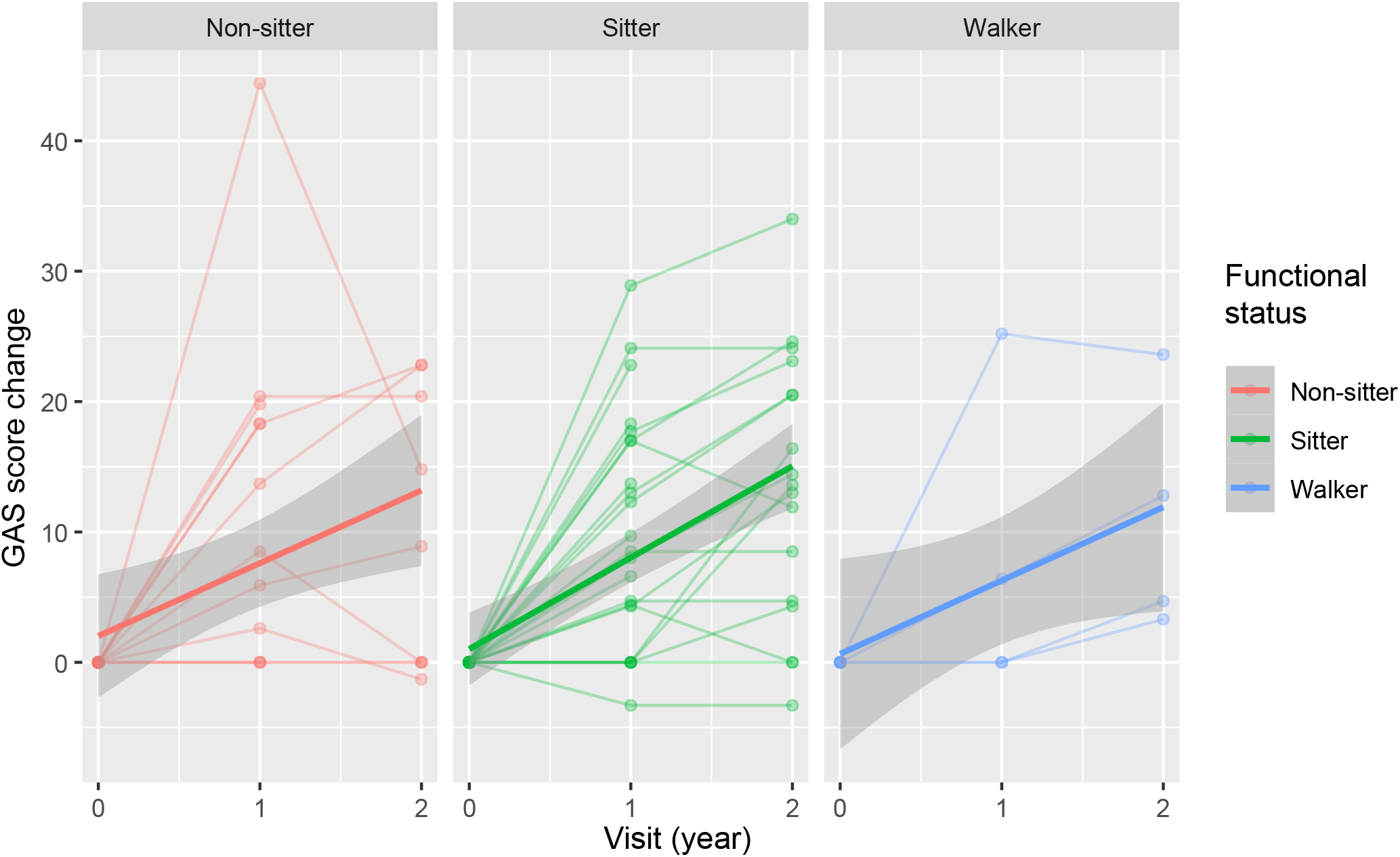
Longitudinal evolution of GAS light change across the two-year follow-up, stratified by baseline functional status. Boxplots displaying the distribution of GAS light change scores at baseline (Year 0), at the 12-month follow-up (Year 1), and at the 24-month follow-up (Year 2), separately for non-sitters (n=14 at baseline), sitters (n=25 at baseline), and walkers (n=5 at baseline). **Abbreviations:** GAS, Goal Attainment Scale

Moreover, at both follow-up visits, patients rated as “minimally improved” on the CGIC showed substantially higher GAS light changes than those rated as “unchanged” or “minimally worse” (Supplementary Figure 4), providing evidence of convergent construct validity from the clinician’s perspective.

The minimal detectable change (MDC) of the GAS light calculated for our cohort was 10.65 points (red line, Supplementary Figure 4). Above this threshold, a change in the GAS light score can be considered to exceed measurement error with 95% confidence.

The MCIC of improvement, obtained by ROC analysis using the CGIC as anchor, was 6.5 points. This value represents the threshold above which GAS light score change can be considered clinically meaningful. The MCIC for worsening could not be calculated, given the low number of patients showing worsening in CGIC.

Using the MCIC threshold, 56.8% of patients (n = 21) at 12 months and 64.5% (n = 20) at 24 months, showed clinically meaningful improvements in their goals, with similar proportions of responders in the different functional subgroups (Table 2). Only one patient at 12 months and 2 patients at 24 months showed deterioration in their goals, all below the -6.5 point threshold and therefore classified as “no change” in table 2 (Figure 4).

**Table 2.**
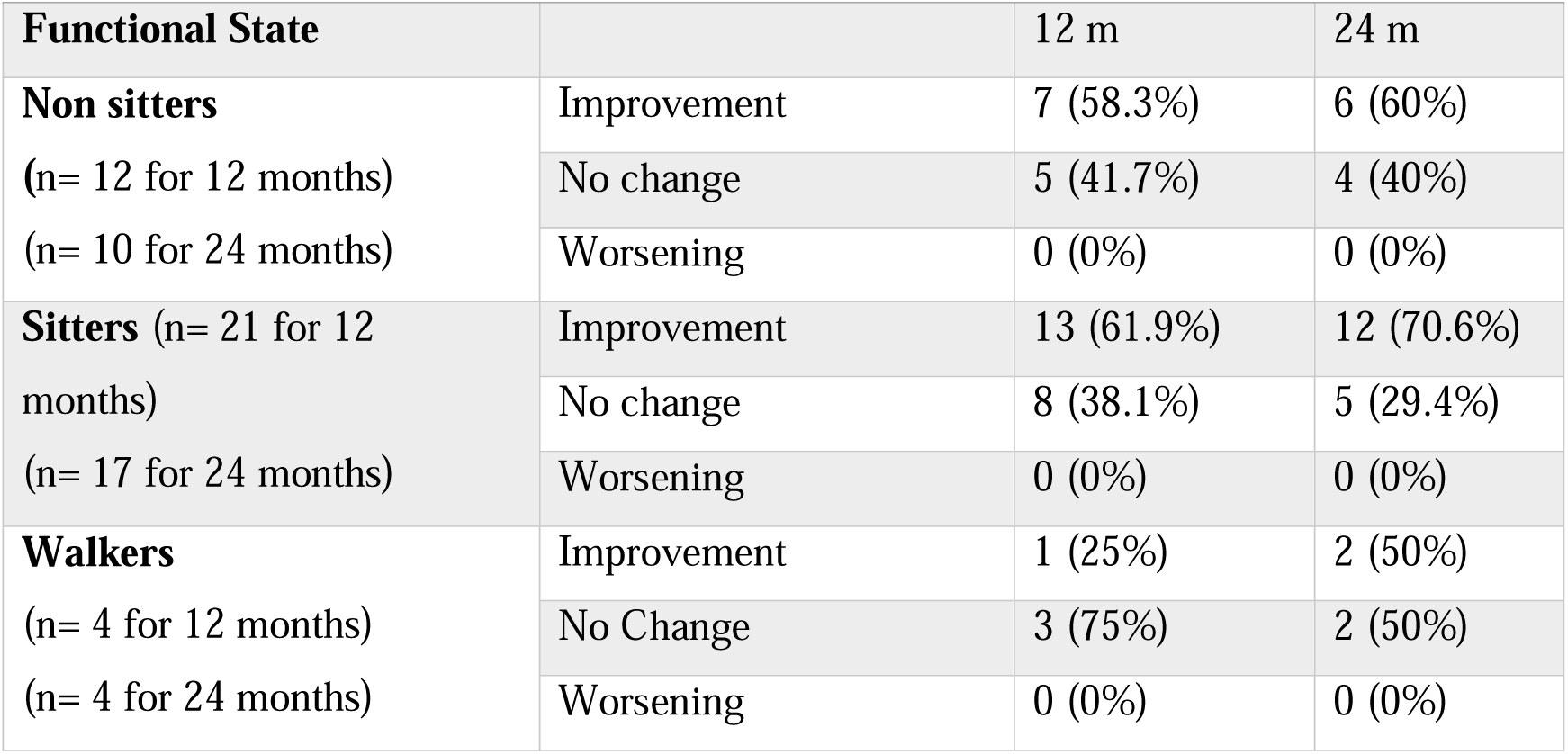
Clinically meaningful GAS light change at 12 and 24 months, stratified by baseline functional status. Values represent the number (percentage) of patients within each functional group categorised by the magnitude of GAS light change at 12 and 24 months relative to baseline. Categories were defined according to the minimal clinically important change (MCIC) threshold (6.5 points) for improvement established in this cohort: *better*, GAS light change ≥+6.5 points; *no change*, GAS light change between −6.5 and +6.5 points; *worse*, GAS light change ≤− 6.5 points. The MCIC for deterioration could not be calculated but was extrapolated to be − 6.5 points for the purposes of this analysis. The analysis includes only patients with complete GAS light data at the 12-month follow-up visit (n=37) and 24-month follow-up visit (n=31). **Abbreviations**: GAS, Goal Attainment Scale; MCIC, minimal clinically important change.

## DISCUSSION

SMA is characterised by an acute or subacute neurodegeneration during the first years of life, followed by a chronic phase of slowly progressive motor decline or stability after adolescence. Beyond primary motor function impairment, long-standing disease is frequently accompanied by multisystem complications affecting musculoskeletal, respiratory, nutritional, and communication domains, which significantly influence autonomy and quality of life. DMTs have dramatically changed the prognosis of SMA patients, with efficacy being highly dependent on SMN2 copy number and, especially, on the delay of treatment since the onset of symptoms. In adults, even when motor gains are frequently not observed, increasing evidence supports the role of DMT in stabilising or slowing disease progression and preserving functional independence[3, 4, 11, 19, 25–27].

While not all adult patients seem to respond equally and several studies have shown larger improvements in younger patients with better motor function [3, 10, 27–30], it remains unclear whether this is the result of the limitations of the motor outcome measures. Given the heterogeneity of adult SMA patients, the selection of appropriate outcome measures to evaluate treatment response is crucial. In clinical practice, motor scales, originally developed or validated in paediatric populations, have been widely adopted for adults, although they show important limitations in some subgroups of patients, including floor and ceiling effects and limited responsiveness to subtle yet clinically meaningful change[5]. As a result, they may underestimate therapeutic benefit in adult patients. To minimise these limitations, the SMA-FCR (a composite score of 6MWT, HFMSE and RULM) was recently validated, showing more sensitivity to change[4]. Despite this, it only detected mild non-statistically significant changes in our study.

Quantitative assessment of muscle strength provides objective, continuous measurements that may overcome some of the floor and ceiling effects observed with ordinal motor scales. Devices such as MyoGrip, MyoPinch or Neuromyotype have been successfully validated and shown to be sensitive in detecting changes in adult SMA patients[31–33]. From a psychometric perspective, pinch strength appears particularly relevant in adult SMA, as it declined significantly over 24 months and showed greater responsiveness than several other quantitative strength measures[33]. Despite this, very few studies have addressed pinch strength in SMA patients after DMT[34], finding no improvements. Our study confirmed those results and found no changes in MyoPinch after risdiplam onset, except a mild non-statistically significant improvement in walkers.

Motor outcomes do not capture bulbar or respiratory function, which may be captured by functional scales such as EK2 and ALSFRS-R. Both show strong construct validity in SMA with little or no floor effect in the non-ambulant population[5]. However, EK2 cannot be used in walkers, while ALSFRS-R, originally developed to evaluate amyotrophic lateral sclerosis, includes items not relevant to SMA[9, 12]. Moreover, both scales show only modest responsiveness to change in adult SMA patients[5].

Accordingly, they were only able to detect mild changes in non-sitter patients in our study.

Patient-reported outcome measures (PROMs) represent a complementary approach that may better reflect real-world response by capturing perceived changes in both motor and non-motor domains such as bulbar or respiratory symptoms, or fatigability, that are frequent and relevant in SMA patients[15, 35]. The GAS light fulfils several criteria that make it particularly suitable for adults with SMA. GAS light allows patients, in collaboration with clinicians, to define individualised treatment goals using the SMART framework. Importantly, the structured quantification of goal importance, probability of achievement, and subsequent attainment enable objective longitudinal analysis while preserving personalisation. Furthermore, GAS light can be applied across the full functional spectrum, from patients with severe motor impairment to ambulant individuals with fewer physical limitations[17, 18, 23]. Interestingly, GAS has been shown to be reliable in SMA patients, while assessing items not captured by conventional motor outcomes[18].

In the present study, we examined three key aspects of GAS light to determine its utility in clinical practice: first, the characterisation of goal types according to functional status; second, its psychometric properties; and third, its ability to detect changes compared with conventional motor and functional scales.

As previously found [21], the goals established by patients were clearly shaped by residual functional capacity and daily life priorities, with goals preferentially targeting areas perceived as modifiable, rather than attempting to recover fully lost abilities.

Remarkably, most goals aimed at improving function, rather than merely at stabilisation. In a disease historically characterised by inexorable progression, functional stability under treatment may represent a major therapeutic achievement. However, the clinical reading of stability or change is therefore inseparable from the natural history of the individual patient, their psychosocial context, and their functional status at treatment initiation, factors that should always inform the interpretation of any outcome measure, including the GAS light. Our results confirm that GAS light is a useful instrument to set personalised goals that are meaningful for patients, but feasible for clinicians too. Non-sitters primarily focused on respiratory function, aiming to preserve ventilatory capacity, improve secretion management, and reduce infections and hospitalisations. Maintenance of upper limb function was also critical to preserve independence in feeding and mobility (e.g., wheelchair use). Sitters demonstrated broader functional concerns. Those with greater impairment defined goals like non-sitters, whereas higher-functioning sitters prioritised upper limb performance and maintenance of standing ability when preserved. Ambulatory patients were more homogeneous, with goals primarily aimed to maintain or improve walking ability and reduce fatigue, a symptom that significantly limits daily activities.

Longitudinal follow-up demonstrated that GAS light captured both clinically meaningful and statistically significant changes across all functional groups after risdiplam treatment. Remarkably, the improvement in GAS light increased with time up to 24 months and showed the same magnitude in all three functional groups. In contrast, conventional motor outcome measures only detected mild non-significant changes in walkers, while functional measures captured changes in non-sitters. In sitters, none of the routinely used scales reflected the improvements found with GAS light.

These findings are in line with previous studies showing stability or mild improvements in motor scales but improvements in PROs after risdiplam treatment, particularly in non-ambulant adults with SMA, where motor scales lack sensitivity [11, 19, 26, 30, 34, 36]. Our findings are also consistent with a recent study where GAS was found more sensitive in detecting improvement in adult SMA patients treated with nusinersen or risdiplam than the standard MFM32[21]. Remarkably, in that study, changes in the Canadian Occupational Performance Measure satisfaction scores correlated with GAS but not with MFM32, highlighting a dissociation between patient-perceived benefit and conventional motor scales, particularly in severely affected patients[21].

Despite its advantages, the use of GAS light in clinical practice requires a psychometric validation. Previous studies have shown its reliability and sensitivity to change [18, 20]. Here, we showed that the distribution of GAS light changes across the CGIC showed an ordered and coherent gradation, providing evidence of convergent construct validity.

Moreover, to facilitate the interpretation of GAS light scores, we calculated the MCIC and MDC. The MCIC was identified at 6.5 points, but the psychometric reliability of the instrument places the MDC at 10.65 points. Here, using the MCIC threshold (6.5 points) we found that 64.5% of patients had reached a clinically meaningful improvement in GAS light scores at 24 months. This is remarkably similar to the 58% of patients who were rated as slightly improved in the CGIC in our study, or the 62.5% of patients achieving at least one of their therapeutic goals found in another study[20], suggesting the internal and external reproducibility of the results. Moreover, when considered alongside the favourable safety profile and high treatment adherence previously reported in this cohort[37], these findings further support risdiplam as a valuable therapeutic option for adults with SMA.

However, it should be noted that GAS light changes falling between the MCIC (6.5) and the MDC (10.65) may reflect measurement variability of the scale rather than a true meaningful change. Therefore, for a more conservative approach, a threshold closer to 10 points could also be considered. Although the establishment of these thresholds requires careful interpretation, given the limited sample size and possible differences in functional subgroups, it offers a frame for the interpretation. Notably, although the MCIC for deterioration could not be calculated due to the limited sample size, only two patients showed mild deterioration in their GAS light scores, both below the MDC and the MCIC for improvement.

Regardless of the threshold chosen, in a population characterised by heterogeneous functional status and slow disease progression, GAS light captured individualised changes that were not consistently detected by conventional motor or functional scales. The concordance observed between GAS light, clinician assessment, and patient-reported global impression further reinforces its validity in routine practice. While fixed-item instruments remain essential for standardised evaluation, our results suggest that integrating individualised, patient-prioritised outcomes may provide a more comprehensive assessment of therapeutic benefit in adult SMA.

### Strengths and limitations

Major strengths of this study include: its population-based design, which makes it representative of the broader adult SMA population, avoiding selection bias; the comprehensive battery of outcome measures, combining motor, functional, and strength scales with a personalised instrument; and the strong statistical approach. Moreover, this study extends previous findings on GAS light by stratifying response across the full functional spectrum, modelling two-year trajectories with linear mixed-effects models, and deriving population-specific MCIC and MDC thresholds anchored to the CGIC, providing, to our knowledge, the first interpretive framework for GAS light scores in adult SMA.

However, the study also has some limitations. First, the limited sample size, particularly within functional subgroups, precluded a stratification of the MDC and MCIC and limited statistical power to detect subgroup-specific effects. Thus, thresholds should be reassessed in future studies with larger cohorts, where cohort-specific reliability estimates can be derived rather than borrowed from the general GAS light literature, and where MDC and MCIC values can be stratified by functional status.

Second, as a real-world, single-arm cohort without an untreated comparator, the study cannot fully exclude the specific effect of risdiplam from the effect of natural history, particularly for goals related to stabilisation.

Third, incomplete follow-up of some patients may have introduced bias, potentially affecting the generalizability of longitudinal findings. However, only 2 patients discontinued risdiplam throughout the study.

Fourth, GAS light itself has intrinsic methodological limitations that restrict its use as a long-term monitoring tool. Its individualised and goal-driven nature makes it most appropriate for point-in-time assessment in the context of a specific intervention or a defined clinical question, such as the evaluation of short- and medium-term (24 months) response after treatment initiation, dose adjustment, or therapeutic switch. It is not designed —nor psychometrically suited— to serve as a stand-alone instrument for continuous long-term follow-up, where goal redefinition, regression to the mean, and the accumulation of measurement error across repeated administrations may compromise interpretability. For long-term disease monitoring, other established motor and functional scales that provide reliable longitudinal trajectories are needed.

Fifth, by relying on individualised goal setting, GAS light introduces potential variability related to the goal-definition process. Despite the use of the SMART framework and a standardised scoring worksheet, differences in clinician facilitation and patient expectations may influence results. Furthermore, expectancy bias cannot be excluded, particularly in an open-label therapeutic context.

## CONCLUSION

The GAS light is a valid, responsive, and clinically interpretable instrument for outcome evaluation in adults with SMA receiving DMT. By incorporating individualised, patient-prioritised goals, GAS light captures dimensions of change that are meaningful for patients yet frequently missed by fixed-item instruments. By providing population-specific interpretive thresholds, we show that GAS light, used alongside conventional scales, helps to detect clinically meaningful changes across the entire functional spectrum of adult SMA patients treated with risdiplam.

## Supporting information

Supplementary Fig 1 Baseline performance

Supplementary Fig 2 Probabilities

Supplementary Fig 3 CGIC-PGIC

Supplementary Fig 4 (CGIC-GAS light)

## DECLARATIONS

### Funding

This study has received funding from CUIDAME (PIC188-18) and from Conselleria de Educación Cultura y Universidades (CIAPOT/2023/25).

The Centro de Investigación Biomédica en Red de Enfermedades Raras (CIBERER) is an initiative from the ISCIII. TS and JFVC are members of the European Reference Network for Rare Neuromuscular Diseases (ERN EURO-NMD).

Sponsors did not participate in the study design, data acquisition and analysis, data interpretation or in writing the article.

### Ethics approval

The study was approved by the Ethics Committee for Biomedical Research of La Fe Hospital (2019-027-1) and was performed in accordance with the 1964 Declaration of Helsinki and its later amendments.

### Consent to participate

Written informed consent was obtained from all individual participants included in the study, as part of their enrolment in the CUIDAME registry (NCT07231549).

### Data availability

JFVC and NCÑG had full access to the database used to create the study population. All data supporting our findings are available on reasonable request.

### Author contributions

NCÑG and KAG participated in clinical data acquisition and interpretation and wrote and edited the manuscript. IPC and TS critically revised the manuscript. DH participated in clinical data interpretation and critically revised the manuscript. JFVC designed the study, participated in clinical data acquisition and interpretation, and wrote and edited the manuscript.

## Acknowledgments

We would like to thank Fernando Mora and Carmen Baviera for their contribution in patients’ assessment, and SMA patients for their participation in the study.

## ABBREVIATIONS

| Abbreviation/Term | Explanation |
| --- | --- |
| 6MWT | 6-Minute Walk Test |
| ALSFRS-R | Amyotrophic Lateral Sclerosis Functional Rating Scale-Revised |
| COPM | Canadian Occupational Performance Measure |
| CGIC | Clinician-Global Impression of Change scale |
| DMTs | Disease-modifying therapies |
| EK2 | Egen Klassifikation 2 |
| GAS light | Goal Attainment Scale light |
| HFMSE | Hammersmith Functional Motor Scale Expanded |
| IQR | Interquartile range |
| kg | Kilograms force |
| MCIC | Minimal Clinically Important Change |
| MDC | Minimal Detectable Change |
| MFM32 | Motor Function Measure - 32 items scale |
| NIV | Non-invasive ventilation |
| PGIC | Patient-Global Impression of Change scale |
| PROMs | Patient-reported outcome measures |
| ROC | Receiver Operating Characteristic |
| RULM | Revised Upper Limb Module |
| SMA | Spinal muscular atrophy |
| SMA-FCR | SMA Functional Composite Score Revised |
| SD | Standard deviation |
| SMN | Survival motor neuron |

## REFERENCES

1. Yu F, Pua DK, Zaita B, et al (2026) Spinal Muscular Atrophy: Advances in Diagnosis, Treatment, and Emerging Therapies. Curr Treat Options Neurol 28:2-. 10.1007/S11940-026-00863-Z

2. Walter MC, Chiriboga C, Duong T, et al (2021) Improving Care and Empowering Adults Living with SMA: A Call to Action in the New Treatment Era. J Neuromuscul Dis 8:543–551. 10.3233/JND-200611

3. Aragon-Gawinska K, Nungo-Garzon NC, Muelas N, et al (2026) Long-term persistence, safety and effectiveness of nusinersen in spinal muscular atrophy: a population-based study. J Neurol 273:430-. 10.1007/S00415-026-13965-0

4. Pasternak A, McDermott MP, Montes J, et al (2025) Spinal Muscular Atrophy Functional Composite Score Revised (SMA-FCR) in Untreated and Nusinersen-Treated Patient Cohorts. Neurology 105:e213839. 10.1212/WNL.0000000000213839

5. Vázquez-Costa JF, Povedano M, Nascimiento-Osorio AE, et al (2022) Validation of motor and functional scales for the evaluation of adult patients with 5q spinal muscular atrophy. Eur J Neurol 29:3666–3675. 10.1111/ene.15542

6. Wu JW, Pepler L, Maturi B, et al (2022) Systematic Review of Motor Function Scales and Patient-Reported Outcomes in Spinal Muscular Atrophy. Am J Phys Med Rehabil 101:590–608. 10.1097/PHM.0000000000001869

7. Pierzchlewicz K, Kepa I, Podogrodzki J, Kotulska K (2021) Spinal Muscular Atrophy: The Use of Functional Motor Scales in the Era of Disease-Modifying Treatment. Child Neurol Open 8:2329048X211008725. 10.1177/2329048X211008725

8. Coratti G, Bovis F, Pera MC, et al (2024) Long-term natural history in type II and III spinal muscular atrophy: a 4-year international study on the Hammersmith Functional Motor Scale Expanded. Eur J Neurol 00:e16517. 10.1111/ENE.16517

9. Brakemeier S, Stolte B, Thimm A, et al (2021) Assessment of Bulbar Function in Adult Patients with 5q-SMA Type 2 and 3 under Treatment with Nusinersen. Brain Sci 11 (9):1244. 10.3390/brainsci11091244

10. Vázquez-Costa JF, Povedano M, Nascimiento-Osorio AE, et al (2022) Nusinersen in adult patients with 5q spinal muscular atrophy: A multicenter observational cohorts’ study. Eur J Neurol 29:3337–3346. 10.1111/ene.15501

11. Brakemeier S, Lipka J, Schlag M, et al (2024) Risdiplam improves subjective swallowing quality in non-ambulatory adult patients with 5q-spinal muscular atrophy despite advanced motor impairment. J Neurol 271: 2649–2657. 10.1007/S00415-024-12203-9

12. Fagoaga J, Girabent-Farrés M, Bagur-Calafat C, et al (2015) Functional assessment for people unable to walk due to spinal muscular atrophy and Duchenne muscular dystrophy. Translation and validation of the Egen Klassifikation 2 scale for the Spanish population. Rev Neurol 60:439–446. 10.33588/RN.6010.2015007

13. Cattinari MG, Pascual-Pascual SI, de Lemus M, et al (2025) Preliminary psychometric validation of patient-reported outcomes relevant to individuals with spinal muscular atrophy and their caregivers. Orphanet J Rare Dis 20:274. 10.1186/S13023-025-03832-Y

14. Slayter J, Casey L, O’Connell C (2023) Patient Reported Outcome Measures in Adult Spinal Muscular Atrophy: A Scoping Review and Graphical Visualization of the Evidence. J Neuromuscul Dis 10:239–250. 10.3233/JND-221595

15. Vázquez-Costa JF, Branas-Pampillón M, Medina-Cantillo J, et al (2023) Validation of a Set of Instruments to Assess Patient- and Caregiver-Oriented Measurements in Spinal Muscular Atrophy: Results of the SMA-TOOL Study. Neurol Ther 12:89–105. 10.1007/S40120-022-00411-2

16. Kiresuk TJ, Smith A, Cardillo JE, eds (1994) Goal Attainment Scaling: Applications, Theory, and Measurement. Routledge, New York. https://www.taylorfrancis.com/books/edit/10.4324/9781315801933/goal-attainment-scaling-thomas-kiresuk-aaron-smith-joseph-cardillo. Accessed 21 Dec 2025

17. Hurn J, Kneebone I, Cropley M (2006) Goal setting as an outcome measure: A systematic review. Clin Rehabil 20:756–772. 10.1177/0269215506070793

18. Casiraghi J, Lizio A, Beretta M, et al (2026) Exploring treatment expectations and clinical meaningfulness in Spinal Muscular Atrophy using the Goal Attainment Scale. Arch Rehabil Res Clin Transl 100590. 10.1016/j.arrct.2026.100590

19. Ñungo Garzón NC, Pitarch Castellano I, Sevilla T, Vázquez-Costa JF (2023) Risdiplam in non-sitter patients aged 16 years and older with 5q spinal muscular atrophy. Muscle Nerve 67:407–411. 10.1002/mus.27804

20. Food and Drug Administration, Center for Drug Evaluation and Research (2023) Patient-Focused Drug Development: Incorporating Clinical Outcome Assessments into Endpoints For Regulatory Decision-Making Guidance for Industry, Food and Drug Administration Staff, and Other Stakeholders.

21. Cintas P, Pouplin S, Debergé L, et al (2026) Feasibility and usefulness of personalised patient-reported outcome measures in the therapeutic follow-up of adult spinal muscular atrophy patients. Clin Neurol Neurosurg, 267:109467. 10.1016/j.clineuro.2026.109467

22. Schmitt JS, Di Fabio RP (2004) Reliable change and minimum important difference (MID) proportions facilitated group responsiveness comparisons using individual threshold criteria. J Clin Epidemiol 57:1008–1018. 10.1016/j.jclinepi.2004.02.007

23. Pike S, Cusick A, Turner-Stokes L, et al (2024) Comparison of standard goal attainment scaling (GAS) and the GAS-light method for evaluation of goal attainment during neurorehabilitation of the upper limb. Journal of the International Society of Physical and Rehabilitation Medicine 7:15–23. 10.1097/PH9.0000000000000028

24. Coratti G, Bovis F, Pera MC, et al (2024) Determining minimal clinically important differences in the Hammersmith Functional Motor Scale Expanded for untreated spinal muscular atrophy patients: An international study. Eur J Neurol. 10.1111/ENE.16309

25. Neuhoff S, Stolte B, Lipka J, et al (2026) Matched-pair analysis of motor outcomes in adults with spinal muscular atrophy on nusinersen vs. risdiplam. J Neurol 273:55-. 10.1007/S00415-025-13589-W

26. Keritam O, Erdler M, Fasching B, et al (2025) Efficacy and safety of risdiplam in adults with 5q-associated spinal muscular atrophy: a nationwide observational cohort study in Austria. EClinicalMedicine 88:103536. 10.1016/J.ECLINM.2025.103536

27. Alonge P, Urbano G, Gadaleta G (2026) Safety and effectiveness of risdiplam in adults with spinal muscular atrophy: a systematic review. J Neurol 273. 10.1007/s00415-025-13557-4

28. Günther R, Wurster CD, Brakemeier S, et al (2024) Long-term efficacy and safety of nusinersen in adults with 5q spinal muscular atrophy: a prospective European multinational observational study. Lancet Reg Health Eur 39. 10.1016/J.LANEPE.2024.100862

29. Maggi L, Bello L, Bonanno S, et al (2020) Nusinersen safety and effects on motor function in adult spinal muscular atrophy type 2 and 3. J Neurol Neurosurg Psychiatry 91:1166–1174. 10.1136/JNNP-2020-323822

30. Gavriilaki M, Moschou M, Pagiantza M, et al (2025) Risdiplam in Adult Patients With 5q Spinal Muscular Atrophy: A Single-Center Longitudinal Study. Muscle Nerve 71:384–391. 10.1002/MUS.28327

31. Lizandra Cortés P, Poveda Verdú D, Albert Férriz A, et al (2024) Validation of Neuromyotype: A smart keyboard for the evaluation of spinal muscular atrophy patients. Neurologia 39:733–742. 10.1016/j.nrl.2022.05.004

32. Seferian AM, Moraux A, Canal A, et al (2015) Upper Limb Evaluation and One-Year Follow Up of Non-Ambulant Patients with Spinal Muscular Atrophy: An Observational Multicenter Trial. PLoS One 10:e0121799. 10.1371/JOURNAL.PONE.0121799

33. Querin G, Lenglet T, Debs R, et al (2021) Development of new outcome measures for adult SMA type III and IV: a multimodal longitudinal study. J Neurol 268:1792–1802. 10.1007/s00415-020-10332-5

34. Iterbeke L, Claeys KG (2025) Two-year Risdiplam treatment in adults with spinal muscular atrophy: improvements in motor and respiratory function, quality of life and fatigue. Neuromuscular Disorders 52: 105397. 10.1016/j.nmd.2025.105397

35. Domine MC, Cattinari MG, de Lemus M, et al (2022) Physical fatigue and perceived fatigability in adolescents and adults with spinal muscular atrophy: A pilot study. Neurology Perspectives 2:199–208. 10.1016/j.neurop.2022.06.008

36. Jaworek A, Jira K, Allen M, et al (2026) Assessment of safety and efficacy of risdiplam treatment in adults with spinal muscular atrophy. Front Neurol 16:1694037. 10.3389/FNEUR.2025.1694037

37. Chovi-Trull M, Ñungo-Garzón NC, Aragon-Gawinska KA, et al (2026) Adherence, Persistence, and Safety of Risdiplam in Spinal Muscular Atrophy: A Population-Based Cohort Study. Neurol Ther 15:1675–1689. 10.1007/S40120-026-00947-7

