## Supplementary material for "Goal Attainment Scale light captures clinically meaningful changes in Adolescents and Adults with Spinal Muscular Atrophy Treated with Risdiplam"

### Methods

*Motor scales*

Motor function was assessed using the Hammersmith Functional Motor Scale Expanded (HFMSE), the Revised Upper Limb Module (RULM), and the 6-Minute Walk Test (6MWT). The HFMSE consists of 33 items, with a maximum score of 66 points, where higher scores indicate better motor function; it was originally developed for high-functioning SMA type 2 and type 3 patients, mainly sitters and walkers. The RULM includes 20 items, with a maximum score of 37 points, where higher scores indicate better upper-limb function, and has been validated in both ambulant and non-ambulant SMA patients. The 6MWT measures the distance walked over 6 min and is applicable only to ambulant patients. In adult SMA patients, these motor scales have shown good construct validity, with strong correlations between motor and bedside functional measures. However, important floor and ceiling effects must be considered: the HFMSE may show a floor effect in the weakest sitters and a ceiling effect in highly functioning walkers, whereas the RULM may show a floor effect in the weakest patients and a ceiling effect in mildly affected ambulant patients. The HFMSE showed strong discriminative ability between walkers and sitters, while the RULM showed moderate discriminative ability; the 6MWT has been validated in ambulant adult SMA patients, although its responsiveness appears limited and should be interpreted cautiously, particularly in small samples.

*Thumb pinch strength*

Maximal thumb pinch strength was measured in the dominant hand using the MyoPinch device and expressed in kilograms. The MyoPinch is a quantitative dynamometer designed to assess isometric key pinch strength with high accuracy, including in patients with marked weakness. Quantitative muscle testing was performed under standardised conditions, with patients receiving uniform instructions to generate maximal voluntary isometric contractions. Then, the maximum value from reproducible trials was retained as the maximal voluntary isometric contraction. From a psychometric perspective, pinch strength appears particularly relevant in adult SMA, as it declined significantly over 24 months in untreated patients and showed greater responsiveness than several other quantitative strength measures.

*Functional scales*

Functional status was assessed using the Egen Klassifikation 2 (EK2) and the revised Amyotrophic Lateral Sclerosis Functional Rating Scale (ALSFRS-R). The EK2 includes 17 items covering eight domains of daily living for non-ambulant patients (wheelchair use, transfers, trunk mobility, eating, swallowing, breathing, coughing, and fatigue) each scored from 0 to 3, with a maximum score of 51 points and higher scores indicating greater disability. The ALSFRS-R includes 12 items across bulbar, upper-limb, lower-limb, and respiratory domains, each scored from 0 to 4, with a maximum score of 48 points and higher scores indicating better function. In adult SMA, bedside functional scales have shown several practical and psychometric advantages over motor function scales: they are faster and easier to administer, capture a broader range of functional states, include clinically relevant non-motor domains, and showed no apparent floor or ceiling effects in the validation cohort.

**Results**

| **Supplementary Table S1. Description of domains and functional regions in Goal Attainment Scale light (GAS light)** | |
|---|---|
| **Functional Regions** | Functional domains |
| **Axial** | Autonomous mobility (rolling in bed, head control, sitting unaided)<br>Personal hygiene |
| **Bulbar** | Feeding (chew and swallow)<br>Dysarthria (speech) |
| **Respiratory** | Noninvasive Ventilation (NIV)<br>Infections<br>Secretion management<br>Speech<br>Dyspnoea |
| **Upper Limbs** | Communication<br>Feeding<br>Personal hygiene<br>Mobility (wheelchair control, transference, grasping objects)<br>Stamina<br>Leisure |
| **Lower Limbs** | Independent walking<br>Stamina |
| **General** | Stamina |

**Supplementary Table S1. Description of functional regions and domains used to categorise patient-set goals in the Goal Attainment Scale light (GAS light).**

Functional regions correspond to anatomically or functionally defined body areas targeted by patients in their individualised GAS light goals. Within each functional region, goals were further categorised into functional domains representing specific tasks or activities. *Autonomous mobility* refers to self-directed displacement or positional change. *Personal hygiene* refers to personal hygiene activities such as washing, grooming, or dressing. *Feeding* refers to autonomous eating and drinking. *Communication* refers to verbal or written interaction, including the use of communication devices. *Leisure* refers to recreational and social activities. *Stamina* refers to endurance during daily or recreational activities. *Speech and dysarthria* refer to the production and clarity of verbal expression. *Secretion management* refers to airway clearance techniques and management of bronchial secretions. *Non-invasive ventilation (NIV)* refers to goals related to ventilatory support, including duration, comfort, and weaning. *Infections* refers to prevention or management of respiratory infections. *Dyspnoea* refers to subjective breathing difficulty.

**Abbreviations**: GAS, Goal Attainment Scale; NIV, non-invasive ventilation.

**Supplementary Table S2. Distribution of GAS light goals by functional region according to functional status.**

| Region | Domain | Non-sitter (n=14) | Sitter (n=25) | Walker (n=5) | Total (n=44) |
|---|---|---|---|---|---|
| **Upper limbs** | **All** | **13 (92.9%)** | **23 (92%)** | **2 (40.0%)** | **38 (86.4%)** |
| | Feeding | 6 (42.9%) | 11 (44%) | 0 (0.0%) | 17 (38.6%) |
| | Mobility | 6 (42.9%) | 8 (32%) | 2 (40.0%) | 16 (36.4%) |
| | Personal hygiene | 1 (7.1%) | 11 (44%) | 0 (0.0%) | 12 (27.3%) |
| | Communication | 6 (42.9%) | 6 (24.0%) | 0 (0.0%) | 12 (27.3%) |
| | Stamina | 0 (0.0%) | 4 (16%) | 0 (0.0%) | 4 (9.1%) |
| | Leisure | 1 (7.1%) | 2 (8%) | 0 (0.0%) | 3 (6.8%) |
| **Lower limbs** | **All** | **0 (0.0%)** | **6 (24.0%)** | **5 (100.0%)** | **11 (25.0%)** |
| | Independent walking | 0 (0.0%) | 6 (24.0%) | 4 (80.0%) | 10 (22.7%) |
| | Stamina | 0 (0.0%) | 0 (0.0%) | 3 (60.0%) | 3 (6.8%) |
| **Axial** | **All** | **5 (35.7%)** | **9 (36%)** | **0 (0.0%)** | **14 (31.8%)** |
| | Autonomous mobility | 5 (35.7%) | 8 (32%) | 0 (0.0%) | 13 (29.5%) |
| | Personal hygiene | 0 (0.0%) | 1 (4%) | 0 (0.0%) | 1 (2.3%) |
| **Respiratory** | **All** | **11 (78.6%)** | **11 (44%)** | **0 (0.0%)** | **22 (50.0%)** |
| | Secretion management | 5 (35.7%) | 5 (20%) | 0 (0.0%) | 10 (22.7%) |
| | Non-invasive ventilation | 2 (14.3%) | 5 (20%) | 0 (0.0%) | 7 (15.9%) |
| | Infections | 4 (28.6%) | 0 (0%) | 0 (0.0%) | 4 (9.1%) |
| | Speech | 1 (7.1%) | 1 (4%) | 0 (0.0%) | 2 (4.5%) |
| | Dyspnoea | 1 (7.1%) | 0 (0.0%) | 0 (0.0%) | 1 (2.3%) |
| **Bulbar** | **All** | **5 (35.7%)** | **5 (20%)** | **0 (0.0%)** | **10 (22.7%)** |
| | Feeding (chew and swallow) | 4 (28.6%) | 4 (16%) | 0 (0.0%) | 8 (18.2%) |
| | Dysarthria (speech) | 2 (14.3%) | 1 (4%) | 0 (0.0%) | 3 (6.8%) |
| **General Stamina** | | **5 (35.7%)** | **6 (24%)** | **3 (60.0%)** | **14 (31.8%)** |

**Supplementary Table S2. Distribution of GAS light goals by functional region according to functional status.**

Values are expressed as the number (percentage) of patients within each functional group who set at least one goal in the corresponding functional region during the baseline GAS light interview. A total of 157 goals were established across the 44 patients. Percentages refer to the proportion of patients within each functional group, not to the proportion of total goals, to preserve the clinical interpretation of individual prioritisation. **Abbreviations**: GAS, Goal Attainment Scale.

**Supplementary Table 3. Linear mixed-effects model assessing SMA-FCR changes with time after risdiplam treatment, adjusting by age, sex and functional status**

| Variables | Estimate | Lower.95. | Upper.95. | P.value |
|---|---|---|---|---|
| Age | -0.042 | -0.346 | 0.261 | 0.796 |
| Male Sex | -0.751 | -9.306 | 7.804 | 0.87 |
| Sitter | 20.699 | 11.505 | 29.893 | <0.001 |
| Walker | 68.884 | 54.115 | 83.653 | <0.001 |
| visit | 0.183 | -0.379 | 0.747 | 0.529 |
| Sitter: visit | -0.318 | -1.076 | 0.439 | 0.418 |
| Walker: visit | 0.868 | -0.285 | 2.02 | 0.148 |

**Supplementary Table 3. Linear mixed-effects model assessing SMA-FCR changes with time after risdiplam treatment, adjusting by age, sex and functional status.**

Sitter and Walker status were associated with significantly higher baseline SMA-FCR scores than non-sitters ($\beta=20.699$ and $\beta=68.884$, respectively; both $p<0.001$), reflecting the expected functional gradient. Neither visit nor age or sex were independently associated with changes in SMA-FCR, although in walkers (walker:visit) there was a trend towards an improvement with visits.

**Supplementary Table 4. Linear mixed-effects model assessing MyoPinch changes with time after risdiplam treatment, adjusting by age, sex and functional status**

| Variables | Estimate | Lower.95. | Upper.95. | P.value |
|---|---|---|---|---|
| Age | 0.016 | -0.012 | 0.043 | 0.296 |

| | | | | |
|---|---|---|---|---|
| Male Sex | 0.124 | -0.789 | 1.036 | 0.803 |
| Sitter | 1.348 | -0.28 | 2.976 | 0.13 |
| Walker | 4.316 | 2.339 | 6.293 | <0.001 |
| visit | -0.051 | -0.574 | 0.464 | 0.852 |
| Sitter: visit | -0.007 | -0.558 | 0.547 | 0.98 |
| Walker: visit | 0.471 | -0.163 | 1.113 | 0.161 |

**Supplementary Table 4. Linear mixed-effects model assessing MyoPinch changes with time after risdiplam treatment, adjusting by age, sex and functional status.**

Walker status was associated with significantly higher baseline MyoPinch scores than non-sitters (β=4.316, 95% CI: 2.339–6.293, p<0.001). Neither visit nor age or sex were independently associated with changes in pinch strength, although in walkers (walker:visit) there was a trend towards an improvement with visits.

| Supplementary Table 5. Linear mixed-effects model assessing EK2 changes with time after risdiplam treatment, adjusting by age, sex and functional status | | | | |
|---|---|---|---|---|
| **Variables** | **Estimate** | **Lower.95.** | **Upper.95.** | **P.value** |
| Age | -0.031 | -0.163 | 0.101 | 0.655 |
| Male sex | 1.646 | -2.407 | 5.7 | 0.444 |
| Sitter | -17.279 | -21.647 | -12.911 | <0.001 |
| visit | -1.365 | -1.99 | -0.745 | <0.001 |
| Sitter: visit | 1.038 | 0.256 | 1.826 | 0.012 |

**Supplementary Table 5. Linear mixed-effects model assessing EK2 changes with time after risdiplam treatment, adjusting by age, sex and functional status.**

As EK2 is designed for non-ambulant patients, walkers were excluded from this model. Sitter status was associated with significantly lower baseline EK2 scores than non-sitters (β=-17.279, 95% CI: -21.647 to -12.911, p<0.001), reflecting less severe impairment. Visit was significantly associated with EK2 score decrease (β=-1.365, p<0.001), while the sitter:visit interaction increased significantly (β=1.038, p=0.012), indicating a different trajectory in sitters vs in non-sitters suggestive of stability.

**Supplementary Table 6. Linear mixed-effects model assessing ALSFRS-R changes with time after risdiplam treatment, adjusting by age, sex and functional status**

| **Variables** | **Estimate** | **Lower.95.** | **Upper.95.** | **P.value** |
|---|---|---|---|---|
| Age | -0.004 | -0.095 | 0.086 | 0.933 |
| Male sex | -1.684 | -4.499 | 1.129 | 0.265 |
| Sitter | 11.371 | 8.034 | 14.709 | <0.001 |
| Walker | 22.57 | 17.317 | 27.822 | <0.001 |
| visit | 1.079 | 0.431 | 1.734 | 0.002 |
| Sitter: visit | -0.993 | -1.811 | -0.189 | 0.02 |
| Walker: visit | -1.025 | -2.24 | 0.197 | 0.107 |

**Supplementary Table 6. Linear mixed-effects model assessing ALSFRS-R changes with time after risdiplam treatment, adjusting by age, sex and functional status**

Sitter and Walker status were associated with significantly higher baseline ALSFRS-R scores than non-sitters (β=11.371 and β=22.570, respectively; both p<0.001), reflecting the expected functional gradient. Visit was significantly associated with ALSFRS-R improvement in non-sitters (β=1.079, p=0.002), which was reversed in sitters (β=-0.993, p=0.02) and walkers (β=-1.025, p=0.107).

**Supplementary Table 7. Linear mixed-effects model assessing GAS light changes with time after risdiplam treatment, adjusting by age, sex and functional status.**

| **Variables** | **Estimate** | **Lower.95.** | **Upper.95.** | **P.value** |
|---|---|---|---|---|
| Age | -0.047 | -0.164 | 0.069 | 0.449 |
| Male sex | -3.056 | -6.608 | 0.497 | 0.112 |
| Sitter | -2.314 | -9.955 | 5.403 | 0.565 |
| Walker | -1.598 | -13.479 | 10.282 | 0.797 |

| Supplementary Table 7. Linear mixed-effects model assessing GAS light changes with time after risdiplam treatment, adjusting by age, sex and functional status. | | | | |
|---|---|---|---|---|
| **Variables** | **Estimate** | **Lower.95.** | **Upper.95.** | **P.value** |
| visit | 5.773 | 2.833 | 8.751 | <0.001 |
| Sitter: visit | 1.347 | -2.392 | 5.005 | 0.483 |
| Walker: visit | -0.064 | -5.605 | 5.463 | 0.982 |

**Supplementary Table 7. Linear mixed-effects model assessing GAS light changes with time after risdiplam treatment, adjusting by age, sex and functional status.**

Linear mixed-effects model of GAS light score change over time following risdiplam treatment, adjusted for age, sex, and baseline functional status (non-sitter as reference), including functional status × visit interaction terms. Visit was significantly associated with GAS light score (β=5.773, $p<0.001$), while age, sex, functional status, and their interaction with visit showed no significant effect, indicating a consistent rate of improvement across functional subgroups.

| Supplementary Table 8. Evolution of GAS light change at 12 and 24 months relative to baseline, stratified by baseline functional status. | | | |
|---|---|---|---|
| **Functional State** | | 12 m | 24 m |
| **Non-sitters** | Improvement | 6 (50%) | 5 (50%) |
| (n= 12 for 12 months) | No significant change | 6 (50%) | 5 (50%) |
| (n= 10 for 24 months) | Worsening | 0 (0%) | 0 (0%) |
| **Sitters** | Improvement | 9 (42.9%) | 11 (64.7%) |
| (n= 21 for 12 months) | No significant change | 12 (57.1%) | 6 (35.3%) |
| (n= 17 for 24 months) | Worsening | 0 (0%) | 0 (0%) |
| **Walkers** | Improvement | 1 (25%) | 2 (50%) |
| (n= 4 for 12 months) | No significant change | 3 (75%) | 2 (50%) |
| (n= 4 for 24 months) | Worsening | 0 (0%) | 0 (0%) |

**Supplementary Table 8. Evolution of GAS light change at 12 and 24 months relative to baseline, stratified by baseline functional status.**

Values represent the number (percentage) of patients within each functional group categorised by the magnitude of GAS light change at 12 and 24 months relative to baseline. The analysis includes only patients with complete GAS light data at the 12-

month follow-up visit (n=37) and 24-month follow-up visit (n=31). Categories were defined according to the minimal detectable change (MDC) threshold (10.65 points) established in this cohort: *better*, GAS light change ≥+10.65 points; *no change*, GAS light change between −10.65 and +10.65 points; *worse*, GAS light change ≤− 10.65 points.

**Abbreviations**: GAS, Goal Attainment Scale; MDC, minimal detectable change.

**Supplementary Figure S1. Baseline functional level of patient-set GAS light goals across functional regions, stratified by baseline functional status.**

Heatmap displaying the mean baseline functional level assigned by patients to their GAS light goals within each functional region, stratified by functional group (non-sitters, sitters, and walkers). At the baseline GAS light interview, patients rated their initial functional status for each goal on a two-point scale, where **−1 indicates partially preserved function** and **−2 indicates absent function**. For each region within each functional group, the value displayed represents the **mean baseline functional level across all goals formulated in that region**. Empty cells indicate functional regions in which no goals were formulated by patients of that group and are therefore not informative.

**Abbreviations:** GAS, Goal Attainment Scale.

**Supplementary Figure S2. Expected probability of goal achievement at the GAS light baseline interview across functional regions, stratified by baseline functional status.**

Heatmap displaying the mean expected probability of achievement assigned by patients to their GAS light goals within each functional region, stratified by functional group (non-sitters, sitters, and walkers). At the baseline GAS light interview, patients rated the expected probability of achieving each goal on a three-point scale: **1 (doubtful), 2 (possible), and 3 (probable)**. For each region within each functional group, the value displayed represents the **mean expected probability across all goals formulated in that region**. Empty cells indicate functional regions in which no goals were formulated by patients of that group.

**Abbreviations:** GAS, Goal Attainment Scale.

**Supplementary Figure S3. Distribution of Clinician- and Patient-Global Impression of Change across the two-year follow-up, stratified by baseline functional status.**

Bar plots displaying the distribution of (A) the Clinician-Global Impression of Change (CGIC) and (B) the Patient-Global Impression of Change (PGIC) at the 12-month (Visit 1) and 24-month (Visit 2) follow-up visits, separately for non-sitters, sitters, and walkers. CGIC responses were categorised as "a little better," "unchanged," or "a little worse"; PGIC responses additionally included the category "much better." Both measures showed a predominance of perceived improvement or stabilisation across all functional subgroups, with few patients reporting worsening at either timepoint.

**Abbreviations:** CGIC, Clinician-Global Impression of Change; PGIC, Patient-Global Impression of Change.

**Supplementary Figure S4. Boxplots displaying the GAS light change distribution at 24-months follow-up across the categories of the Clinician Global Impression of Change (CGIC).**

Of the seven possible CGIC categories, only three were represented in this cohort ("minimally improved", "unchanged", "minimally worse"); no patient was categorised as "moderately/much improved" or "moderately/much worse" by their treating clinician. The horizontal green line at 6.5 points indicates the minimal clinically important change for the GAS light while the red line at 10.65 indicates the minimal detectable change.

**Abbreviations:** CGIC, Clinician Global Impression of Change; GAS, Goal Attainment Scale; MCIC, minimal clinically important change; MDC, minimal detectable change.
