## Supplementary figures and images for "Goal Attainment Scale light captures clinically meaningful changes in Adolescents and Adults with Spinal Muscular Atrophy Treated with Risdiplam"

### Supplementary Fig 1 Baseline performance

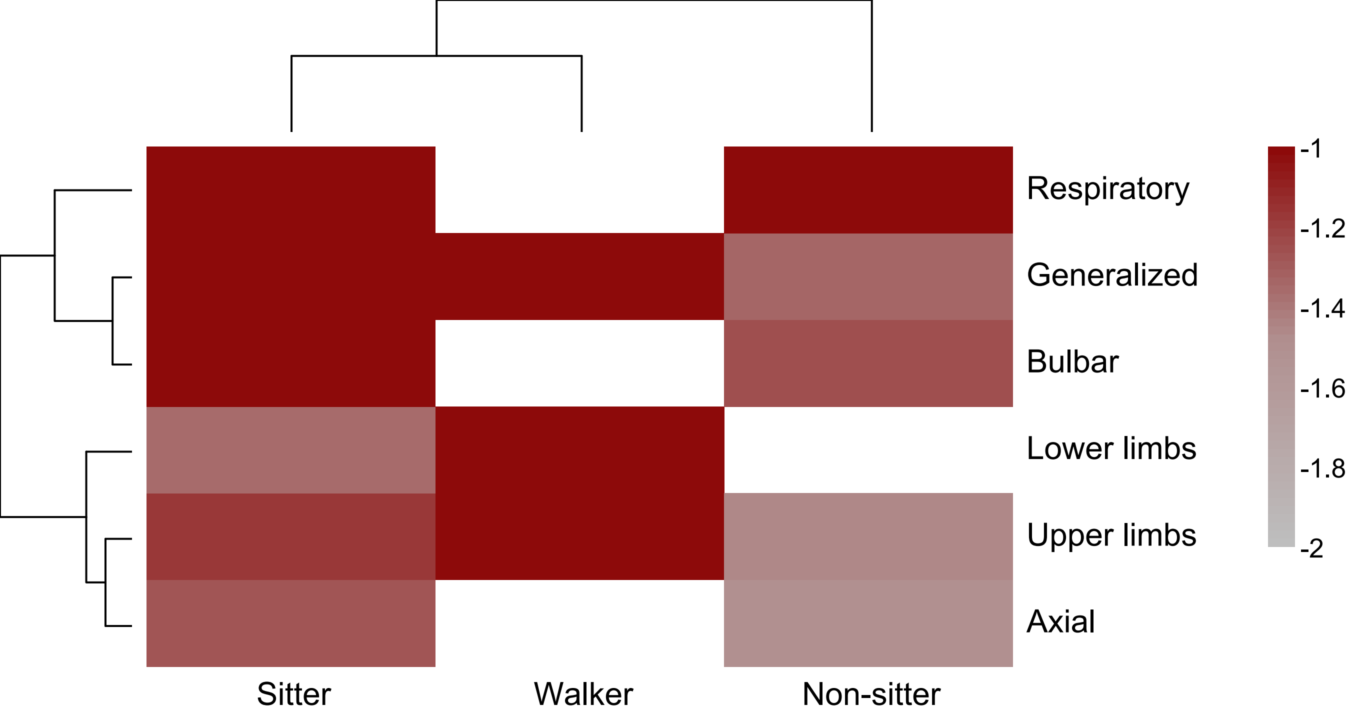

### Supplementary Fig 2 Probabilities

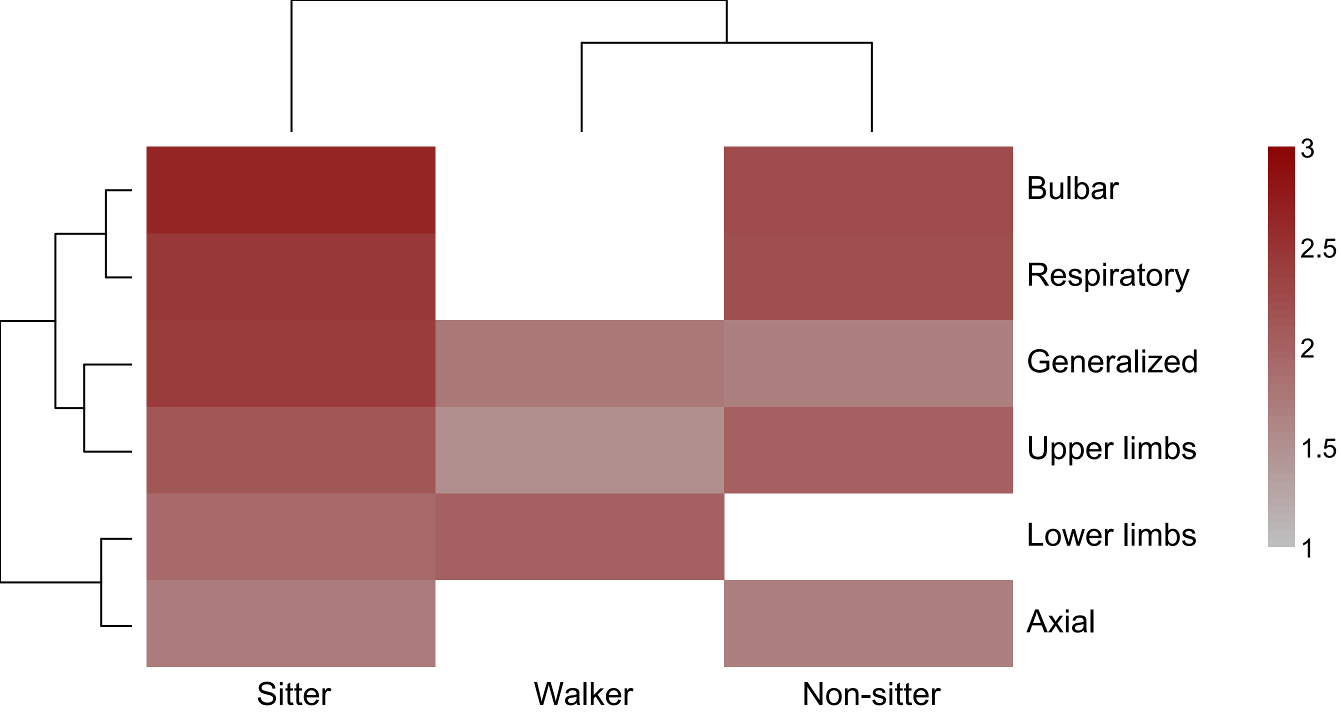

### Supplementary Fig 3 CGIC-PGIC

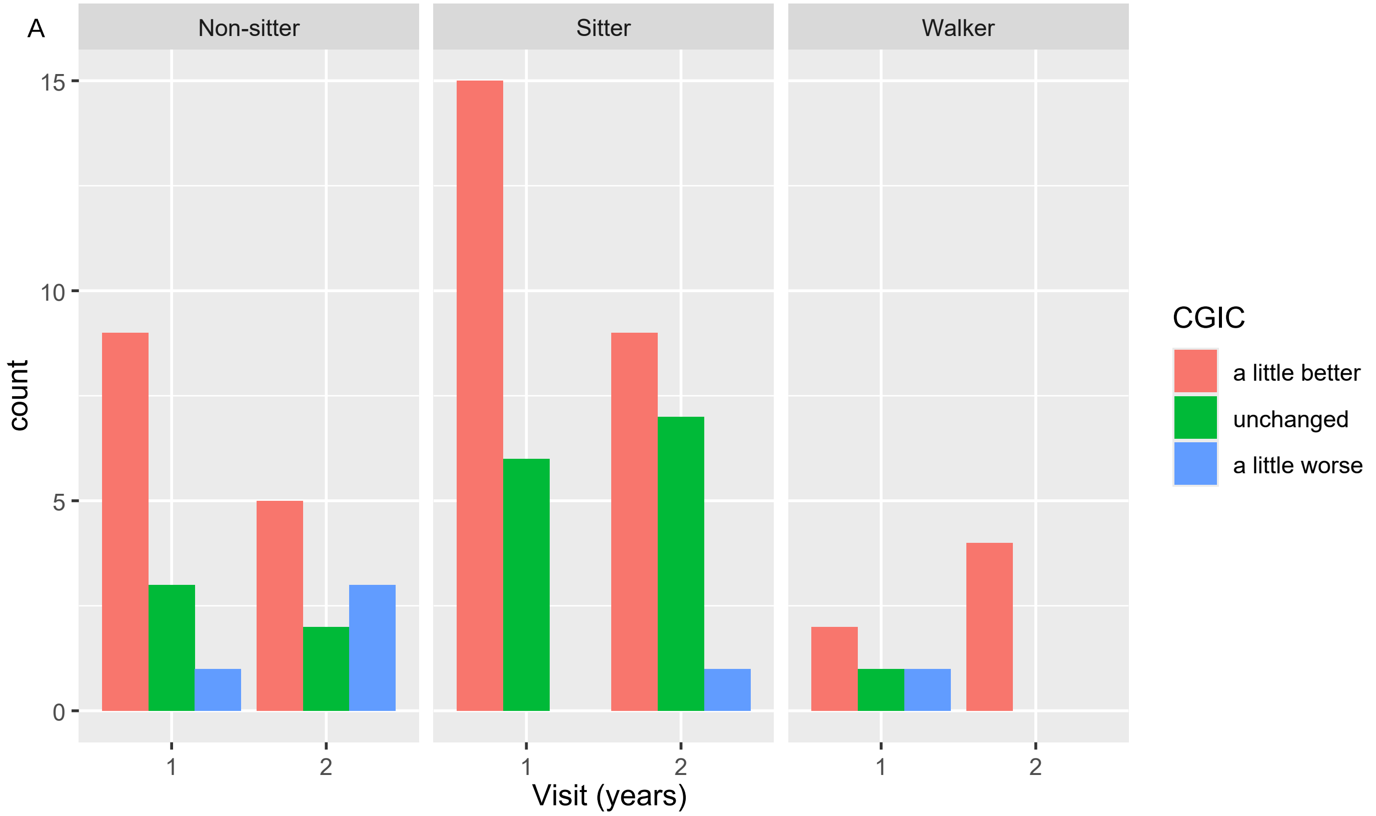

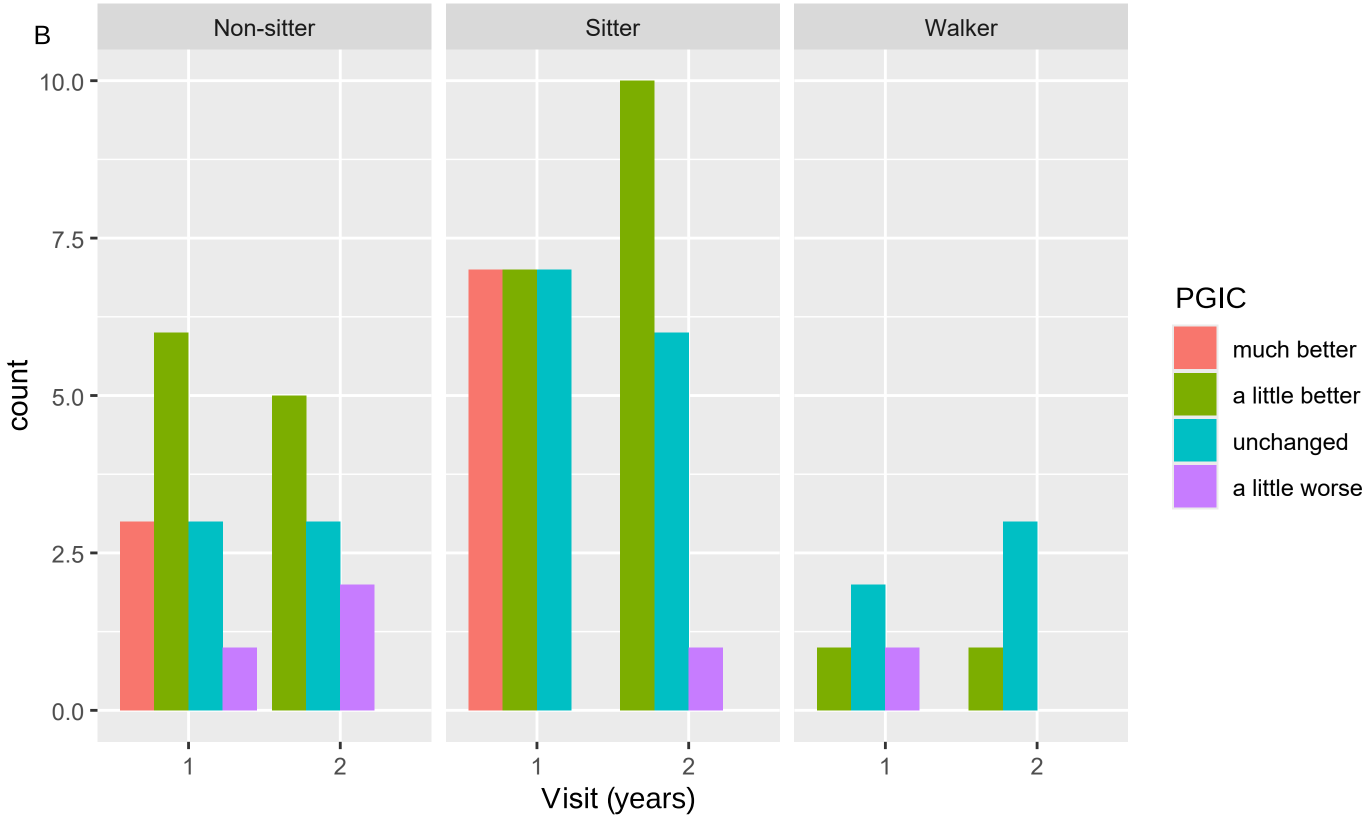

### Supplementary Fig 4 (CGIC-GAS light)

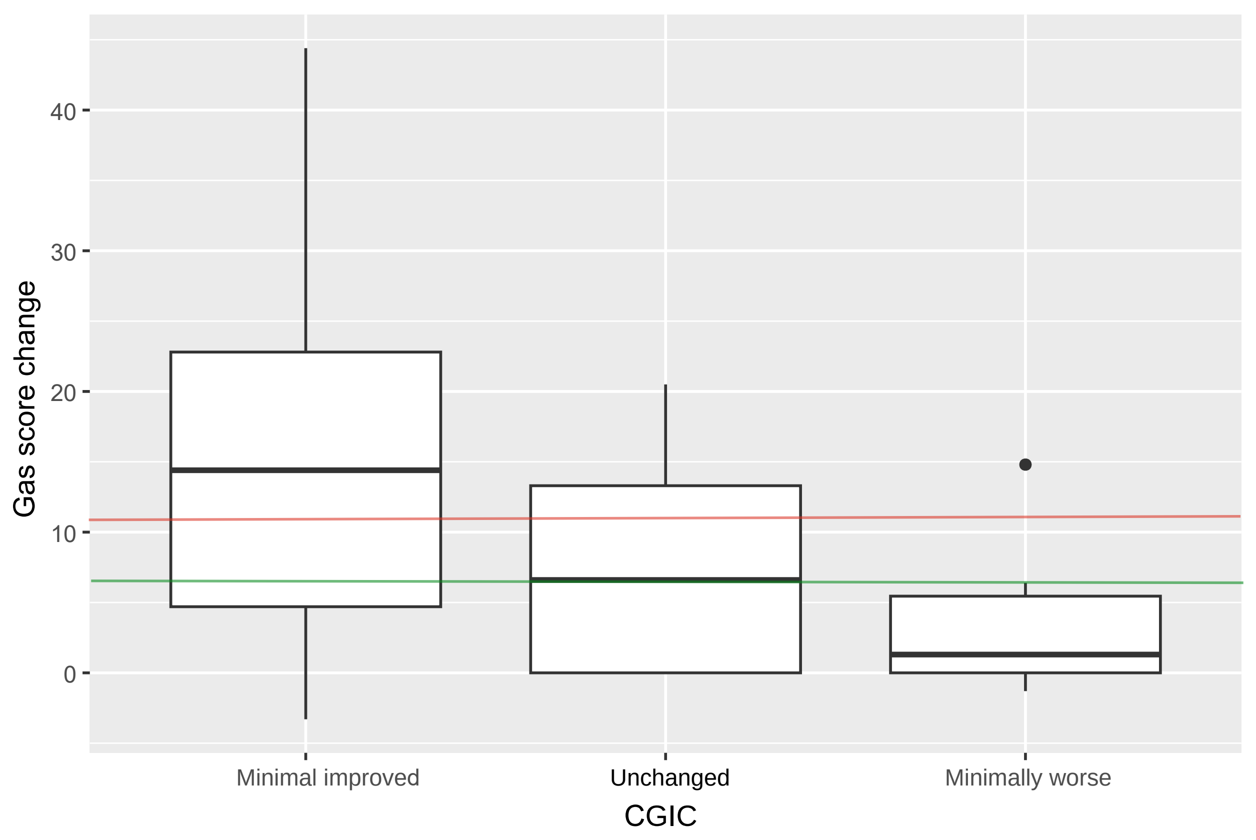
